# Global Burden Of Problematic Internet Use: An Umbrella Review and Metanalysis

**DOI:** 10.64898/2026.05.23.26353953

**Authors:** Tiara Schwarze-Taufiq, Sophie Weber, Blanca Larrain, Gabriel Gatica-Bahamonde, Ornella Corazza, Jessica Neicun, Dan J. Stein, Konstantinos Ioannidis, Zsolt Demetrovics, Sam R. Chamberlain, Lior Carmi, Joseph Zohar, Hans-Jurgen Rumpf, Natalie Hall, Jose M. Menchon, Célia Sales, Christian Montag, Katajun Lindenberg, Mart Susi, Anja Huizink, Marc N. Potenza, Stefano Pallanti, Nicholas Morgan, Carmen Moreno, Diane Purper-Ouakil, Matthias Brand, Murat Yucel, Andrea Czako, Susanne Walitza, Julius Burkauskas, Katalin Felvinczi, Megan Smith, David Wellsted, Julia Jones, Teresa Silva Dias, Simon Foster, Meichun Mohler-Kuo, Ina Neumann, Erica Fongaro, Sara Fally, Hernani Oliveira, Renzo Abregú-Crespo, Marta Sepúlveda-Palomo, Naomi Fineberg, Robin van Kessel, Andres Roman-Urrestarazu

**Affiliations:** Mental Health | Policy | Economics Group, Department of Psychiatry, University of Cambridge, Cambridge, United Kingdom; Centre for Mathematical Modelling of Infectious Diseases, London School of Hygiene and Tropical Medicine, London, United Kingdom; Addiction Division, University Hospitals of Geneva, Geneva, Switzerland; School of Life and Medical Sciences, University of Hertfordshire, Hatfield, United Kingdom; Addiction Science Lab, Department of Psychology and Cognitive Science, University of Trento, Trento, Italy; SAMRC Unit on Risk & Resilience in Mental Disorders, Department of Psychiatry and Neuroscience Institute, University of Cape Town, Cape Town, South Africa; Hampshire and Isle of Wight Healthcare NHS Foundation Trust, Southampton, United Kingdom; Department of Psychiatry, University of Southampton, Southampton, United Kingdom; Institute of Psychology, ELTE Eötvös Loránd University, Budapest, Hungary; Centre of Excellence in Responsible Gaming, University of Gibraltar, Gibraltar; Institute for Mental Health and Wellbeing, College of Education, Psychology and Social Work, Flinders University, Adelaide, Australia; Department of Psychiatry, Faculty of Medicine, University of Southampton and Hampshire and Isle of Wight Healthcare NHS Foundation Trust, Southampton, United Kingdom; Data Science Institution, Reichman University, Herzliya, Israel; Sackler Medical School, Tel Aviv University, and Chaim Sheba Medical Center Tel Hashomer, Tel Aviv, Israel; Department of Psychiatry and Psychotherapy, Translational Psychiatry Unit, University of Lübeck, and Center for Behavioral Addiction Research (CeBAR), University Duisburg-Essen, Germany; Department of Psychiatry, Bellvitge University Hospital-IDIBELL; Department of Clinical Sciences, Faculty of Medicine and Health Sciences, University of Barcelona; CIBERSAM, Spain; Faculty of Psychology and Education Sciences, University of Porto, Porto, Portugal; Centre for Cognitive and Brain Sciences, Institute of Collaborative Innovation, University of Macau, Macau, China; Psychologisches Institut, Universität Heidelberg, Heidelberg, Germany; Tallinn University, Tallinn, Estonia; Faculty of Behavioural and Movement Sciences, Department of Clinical, Neuro- and Developmental Psychology, Vrije Universiteit Amsterdam, Amsterdam, Netherlands; Department of Psychiatry, Yale University School of Medicine, New Haven, CT, USA; Yale Child Study Center, Yale University, New Haven, CT, USA; Connecticut Mental Health Center, New Haven, CT, USA; Connecticut Council on Problem Gambling, Wethersfield, CT, USA; Department of Neuroscience, Yale University, New Haven, CT, USA; Wu Tsai Institute, Yale University, New Haven, CT, USA; Istituto di Neuroscienze, Firenze, Italy; Euro Youth Mental Health CIC; Department of Child and Adolescent Psychiatry, Institute of Psychiatry and Mental Health, Hospital General Universitario Gregorio Marañón, IiSGM, CIBERSAM, ISCIII, School of Medicine, Universidad Complutense, Madrid, Spain; Service de médecine psychologique de l’enfant et de l’adolescent, CHU de Montpellier– Hôpital Saint-Éloi, Université de Montpellier, Montpellier, France; Équipe psychiatrie, développement et trajectoires, Inserm U1018, CESP, Université Paris-Saclay, Villejuif, France; Department of Molecular Psychology, Institute of Psychology and Education, Ulm University, Ulm, Germany; QIMR Berghofer Medical Research Institute, Herston, Queensland, Australia; Department of Child and Adolescent Psychiatry, University of Zurich, Zurich, Switzerland; Neuroscience Center Zurich, Swiss Federal Institute of Technology and University of Zurich, Zurich, Switzerland; Zurich Center for Integrative Human Physiology, University of Zurich, Zurich, Switzerland; Laboratory of Behavioral Medicine, Neuroscience Institute, Lithuanian University of Health Sciences, Palanga, Lithuania; Department of Psychiatry and Behavioral Sciences, University of Minnesota, Minneapolis, MN, USA; Institute of Public Health of the University of Porto (ISPUP); Instituto de Investigación Sanitaria Gregorio Marañón (IiSGM), Madrid, Spain; Department of Clinical, Pharmaceutical and Biological Science, Centre for Health Services and Clinical Research, Cognitive Neuropsychology, School of Life and Medical Sciences, University of Hertfordshire, United Kingdom; Department of International Health, Care and Public Health Research Institute (CAPHRI), Faculty of Health, Medicine and Life Sciences, Maastricht University, Maastricht, Netherlands; LSE Health, Department of Health Policy, London School of Economics and Political Science, London, United Kingdom; Departamento de Psiquiatría, Pontificia Universidad Católica de Chile, Santiago de Chile, Chile

## Abstract

**Importance:** Problematic use of the internet (PUI) behaviors, including problematic gaming, social media use, smartphone use, and general internet use, have been increasingly studied worldwide. So far it is unclear what the global prevalence of PUI is.

**Objective:** To critically appraise existing systematic reviews and meta-analyses on the prevalence of PUI behaviors and generate aggregated global prevalence estimates across different manifestations and definitions.

**Data Sources:** MEDLINE (Ovid), Embase (Ovid), Scopus, Web of Science, CINAHL, and the Cochrane Review Library were searched for relevant articles from database inception to the most recent available search prior to manuscript preparation. Searches targeted systematic reviews and meta-analyses reporting prevalence for PUI-related behaviors.

**Study Selection:** Systematic reviews and meta-analyses of observational studies reporting prevalence estimates for problematic gaming, problematic internet use, problematic smartphone use, problematic social media use, or sexting were included. Scoping reviews were retained for descriptive synthesis only.

**Data Extraction and Synthesis:** An umbrella review methodology was used. Data extraction and methodological appraisal were conducted using AMSTAR-2 to assess the quality of included systematic reviews up to February 2026. Primary studies included in each review were extracted and pooled using random-effects meta-analysis. Analyses were conducted to estimate pooled prevalence with 95% confidence intervals (CIs) and heterogeneity across non-overlapping primary studies. Small-study effects were examined.

**Main Outcomes and Measures:** Global pooled prevalence estimates for PUI behaviors, including problematic gaming, problematic internet use, problematic smartphone use, problematic social media use, and sexting.

**Results:** Eleven reviews (10 systematic reviews and 1 scoping review) met inclusion criteria, representing data from 3 145 428 individuals, of whom 3 030 023 were included in pooled prevalence analyses. Across regions, pooled prevalence estimates were 6% (95% CI, 5%–7%) for problematic gaming, 16% (95% CI, 15%–17%) for problematic internet use, 32% (95% CI, 28%–35%) for problematic smartphone use, and 23% (95% CI, 19%–28%) for problematic social media use. Substantial heterogeneity (I^2^ > 99%) was observed across primary studies, reflecting variation in study methodologies, sampled populations, and definitions of PUI behaviors.

**Conclusions and Relevance:** PUI behaviors appear to affect a substantial proportion of the global population. However, methodological concerns were common, with 9 of 10 systematic reviews rated as having low or critically low confidence according to AMSTAR-2. Evidence remains concentrated in East Asia and Europe, and many reviews combine heterogeneous populations and sampling strategies. Additional high-quality epidemiological research, including underrepresented regions is needed to refine prevalence estimates, clarify risk factors, and support the development of standardized criteria for PUI behaviors.

**Key Points:** *Question:* What is the global prevalence of problematic use of the internet (PUI) behaviors across major subtypes, based on an umbrella review of existing reviews and a meta-analysis of nonoverlapping primary studies?

*Findings:* In this umbrella review of 11 reviews and random-effects meta-analysis of 753 nonoverlapping primary studies including 3 030 023 individuals, pooled prevalence estimates were 6% for problematic gaming, 16% for problematic internet use, 32% for problematic smartphone use, and 23% for problematic social media use. Findings indicated substantial between-study heterogeneity, and most included systematic reviews were rated low or critically low in methodological quality.

*Meaning:* These findings suggest that PUI behaviors affect a substantial proportion of the global population, but more methodologically rigorous and geographically representative epidemiological research is needed to generate more precise and comparable prevalence estimates across clinical and population samples.

## 1. Introduction

The pervasive influence of the internet in contemporary society has reshaped the way individuals interact, work, and navigate the world ^1,2^. While the digital age has ushered in unparalleled connectivity and convenience, it has also brought to light a concerning phenomenon linked to digital exposure, namely problematic use of the internet (PUI) ^3^. Characterised by compulsive or dysfunctional online behaviours such as pathologically high levels of social media engagement, pornography use, gaming, online gambling, and online shopping, PUI has emerged as a pressing public health concern with profound implications for individual well-being across the lifespan, including social and occupational impairment, cognitive dysfunction, poor eye health, and poor mental health ^2,4,5^. While these specific behaviours capture a segment of how PUI can manifest, each of these refers to a specific pervasive behavioural pattern in relation to the internet and fails to recognise the many ways in which internet can be harmful outside of these diagnoses ^6^. PUI can have significant and multifaceted health consequences, impacting various aspects of an individual’s well-being ^1,7^. Nevertheless, the multifaceted and heterogenous nature of PUI has made it a difficult phenomenon to define, diagnose, and treat, with researchers and health professionals calling for a consistent conceptualization for this expansive yet fragmented condition ^3^.

Estimates of PUI prevalence to date are varied, and work is needed to elucidate reasons for this variation. Some estimates reported that up to 27% of the global population reports experiencing some form of PUI, with a disproportionately higher impact observed in lower and lower-middle-income countries ^8^. By contrast, studies conducted among adults in various countries have reported prevalence rates of PUI ranging from 1% to 9%, with differences reflecting heterogeneity in populations studied, definitions used, regions represented, and variety in measurement instruments ^9,10^. One key source of heterogeneity is operationalization of PUI. Foundational work in problematic internet use behaviour by Young et al. in 1992 initially conceptualised PUI as an impulse-control disorder and proposed classification of general internet addiction adapted from problematic gambling criteria.As such, many existing evidence syntheses focus on literature relating to “general internet addiction,” without explicit consideration of the specific online activities involved^11^. Research in the United States has shown that approximately 9.8% of adults exhibit symptoms of internet addiction according to this older conceptualisation ^11^. Similarly, studies in other countries, including China and the Netherlands, have reported prevalence rates ranging from 2% to 9% among adults ^9,10^. As research into problematic digital behaviours has evolved over time, the term PUI was introduced to represent a more expansive, multifaceted constellation of distinct, but interrelated problematic digital behaviours that may or may not qualify as “addictions.” The first empirical publication concerning PUI as a multifaceted umbrella term for diverse internet use behaviours was released in 2018, delineating specific behaviours including social media, online pornography, and gaming^11^. Epidemiological research into PUI is limited by the lack of official criteria (aside from those clinically recognised in ICD-11), leading to fragmented measurement of PUI ^12^. While some attempts have been made to start synthesizing existing research on PUI, no comprehensive review of those existing reviews has been performed to assess whether these evidence syntheses use harmonious or diverging conceptualisations of problematic internet use. As a result, the burden of disease surrounding this condition remains unclear, making it difficult to prioritise it on public policy agendas. This article aims to provide an umbrella synthesis of existing reviews on PUI, with the goal to provide a more comprehensive insight on the burden of disease, as well as to highlight good practices and gaps in research.

## 2. Methods

This umbrella review is performed according to JBI guidelines ^13^, and reported according to the Preferred Reporting Items for Systematic Reviews and Meta-Analyses (PRISMA) 2020 reporting guidelines ^14^. Due to the epidemiological nature of the research question, we structure the eligibility criteria according to the Condition-Context-Population framework ^15^.

### 2.1. Eligibility criteria

To be eligible for inclusion in this umbrella review, existing reviews had to contain a measurement of the prevalence of problematic internet use, internet addiction, social media addiction, video game addiction, gaming disorder, or technology addiction. Reviews covering any geographic region in the world during any time period are considered. This review focusing on participants of any biological sex, racial/ethnic background, health status, socioeconomic status, urbanicity, and geographical location. Participants can be children (aged 0-14), youth (aged 15-24), adults (aged 25-65) or the older population (aged 65 and over) ^16,17^. Systematic reviews with or without meta-analysis and scoping reviewsthat analyze the epidemiology of problematic internet use will be considered for this review. To be eligible, the literature review should follow standardised methodology and should have a clearly articulated search strategy with more than one data source ^18^. Only studies in English were considered to allow the researchers to assess how various subcategories of problematic internet use have been defined and operationalised across the included reviews. Because we planned to undertake quantitative analysis, we deviated from the PROSPERO protocol by excluding narrative and critical reviews.

### 2.2. Search strategy

The databases MEDLINE (Ovid), Embase (Ovid), Scopus, Web of Science, CINAHL, Cochrane Review Library were searched for relevant scientific articles. These databases were chosen to cover both health-specific and interdisciplinary academic fields, which is important for the topic of problematic internet use as this lies at the intersection of health, technology, and social sciences. The full build-up of the query for MEDLINE and Embase are shown in Table 1, while the queries for other databases are included in the eMethods in the appendix. The search string was informed by previous reviews on problematic internet use or internet addiction ^8,19,20^. Two researchers (RVK and ARU) screened the titles and abstracts. Cohen’s K was computed to indicate the inter-rater agreement score. One reviewer (SW) performed the full-text screening, which was validated by a second reviewer (RVK).

**Table 1:**
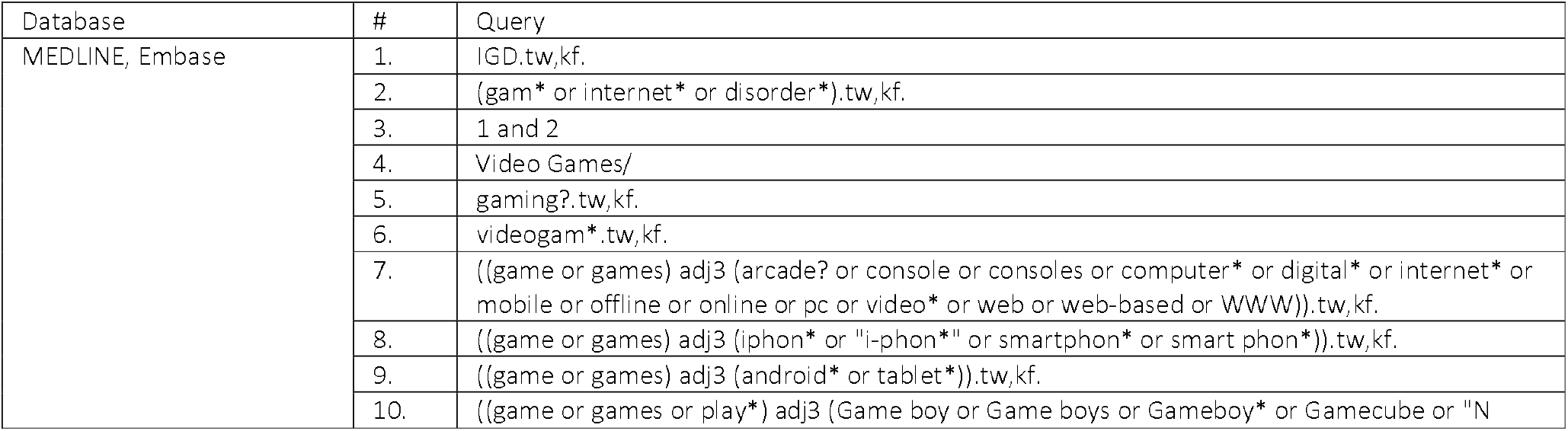
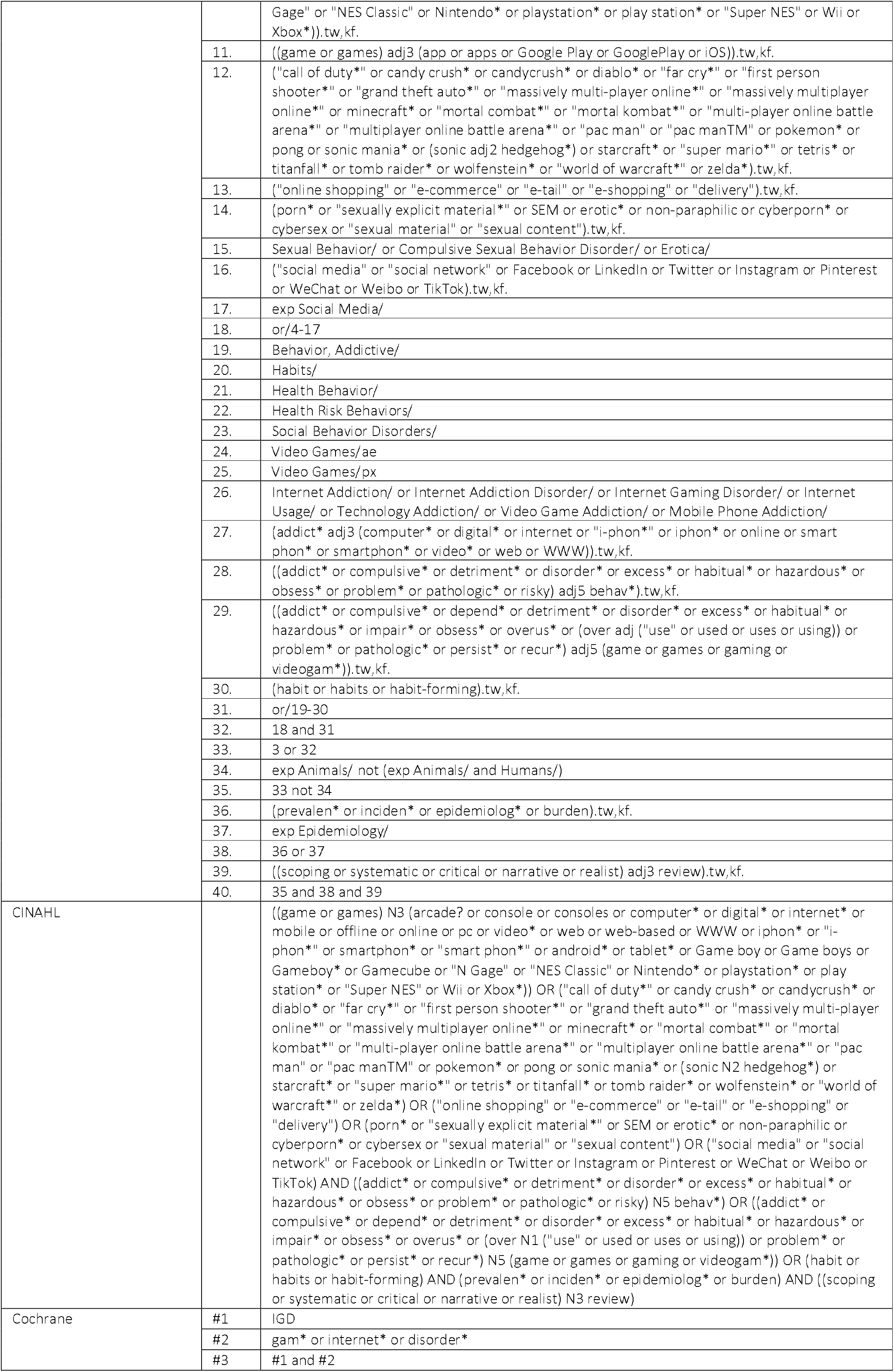
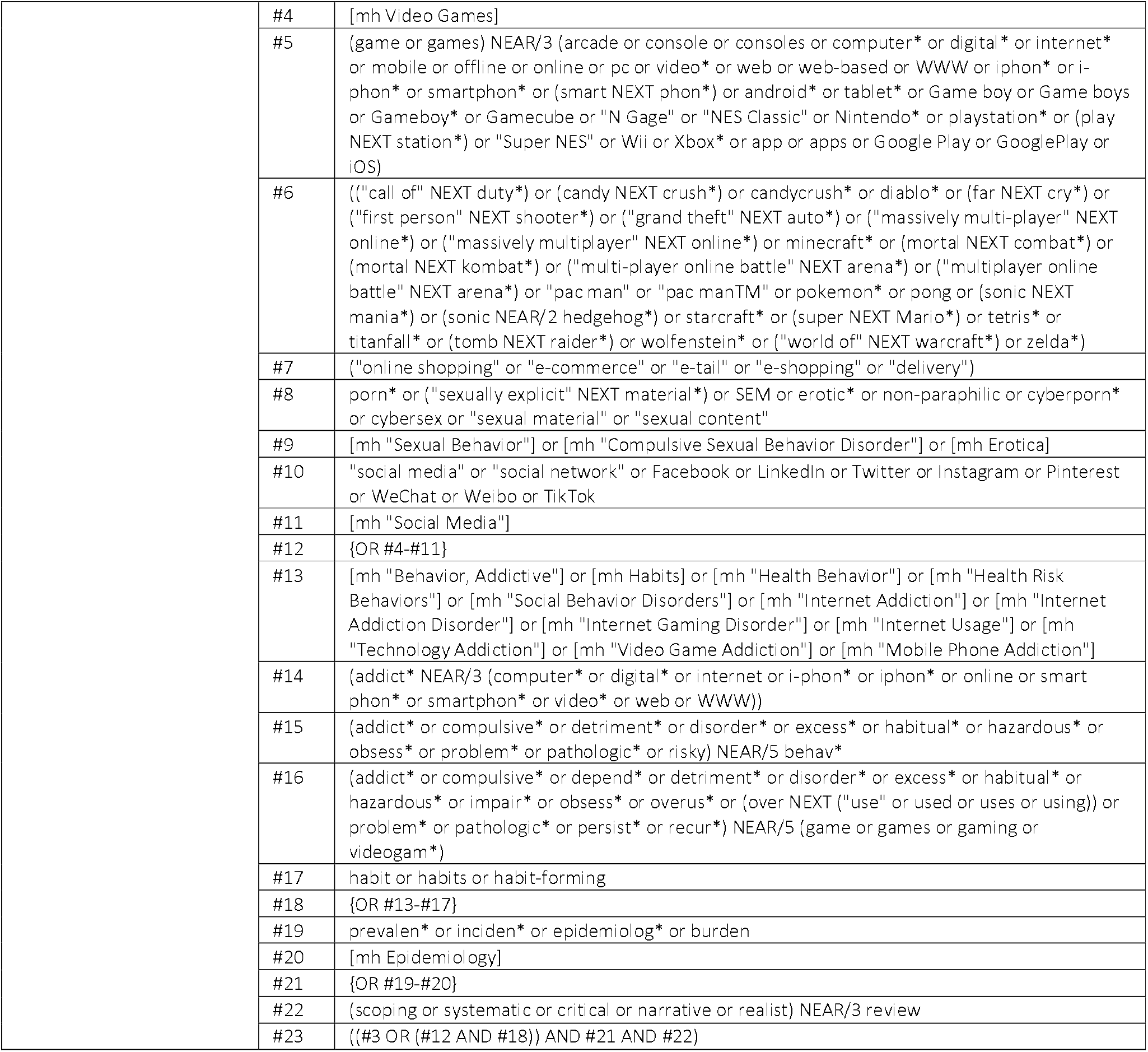
Build-up of the search query for scientific databases in Ovid.

| Database | # | Query |
| --- | --- | --- |
| MEDLINE, Embase | 1. | IGD.tw,kf. |
|  | 2. | (gam* or internet* or disorder*).tw,kf. |
|  | 3. | 1 and 2 |
|  | 4. | Video Games/ |
|  | 5. | gaming?.tw,kf. |
|  | 6. | videogam*.tw,kf. |
|  | 7. | ((game or games) adj3 (arcade? or console or consoles or computer* or digital* or internet* or mobile or offline or online or pc or video* or web or web-based or WWW)).tw,kf. |
|  | 8. | ((game or games) adj3 (iphon* or "i-phon*" or smartphon* or smart phon*)).tw,kf. |
|  | 9. | ((game or games) adj3 (android* or tablet*)).tw,kf. |
|  | 10. | ((game or games or play*) adj3 (Game boy or Game boys or Gameboy* or Gamecube or "N |
|  |  | Gage" or "NES Classic" or Nintendo* or playstation* or play station* or "Super NES" or Wii or Xbox*)),tw,kf. |
|  | 11. | ((game or games) adj3 (app or apps or Google Play or GooglePlay or iOS)).tw,kf. |
|  | 12. | ("call of duty*" or candy crush* or candycrush* or diablo* or "far cry*" or "first person shooter*" or "grand theft auto*" or "massively multi-player online*" or "massively multiplayer online*" or minecraft* or "mortal combat*" or "mortal kombat*" or "multi-player online battle arena*" or "multiplayer online battle arena*" or "pac man" or "pac manTM" or pokemon* or pong or sonic mania* or (sonic adj2 hedgehog*) or starcraft* or "super mario*" or tetris* or titanfall* or tomb raider* or wolfenstein* or "world of warcraft*" or zelda*).tw,kf. |
|  | 13. | ("online shopping" or "e-commerce" or "e-tail" or "e-shopping" or "delivery").tw,kf. |
|  | 14. | (porn* or "sexually explicit material*" or SEM or erotic* or non-paraphilic or cyberporn* or cybersex or "sexual material" or "sexual content").tw,kf. |
|  | 15. | Sexual Behavior/ or Compulsive Sexual Behavior Disorder/ or Erotica/ |
|  | 16. | ("social media" or "social network" or Facebook or LinkedIn or Twitter or Instagram or Pinterest or WeChat or Weibo or TikTok).tw,kf. |
|  | 17. | exp Social Media/ |
|  | 18. | or/4-17 |
|  | 19. | Behavior, Addictive/ |
|  | 20. | Habits/ |
|  | 21. | Health Behavior/ |
|  | 22. | Health Risk Behaviors/ |
|  | 23. | Social Behavior Disorders/ |
|  | 24. | Video Games/ae |
|  | 25. | Video Games/px |
|  | 26. | Internet Addiction/ or Internet Addiction Disorder/ or Internet Gaming Disorder/ or Internet Usage/ or Technology Addiction/ or Video Game Addiction/ or Mobile Phone Addiction/ |
|  | 27. | {addict* adj3 (computer* or digital* or internet or "i-phon*" or iphon* or online or smart phon* or smartphon* or video* or web or WWW)).tw,kf. |
|  | 28. | ((addict* or compulsive* or detriment* or disorder* or excess* or habitual* or hazardous* or obsess* or problem* or pathologic* or risky) adj5 behav*).tw,kf. |
|  | 29. | ((addict* or compulsive* or depend* or detriment* or disorder* or excess* or habitual* or hazardous* or impair* or obsess* or overus* or (over adj ("use" or used or uses or using)) or problem* or pathologic* or persist* or recur*) adj5 (game or games or gaming or videogam*)).tw,kf. |
|  | 30. | (habit or habits or habit-forming).tw,kf. |
|  | 31. | or/19-30 |
|  | 32. | 18 and 31 |
|  | 33. | 3 or 32 |
|  | 34. | exp Animals/ not (exp Animals/ and Humans/) |
|  | 35. | 33 not 34 |
|  | 36. | (prevalen* or inciden* or epidemiolog* or burden).tw,kf. |
|  | 37. | exp Epidemiology/ |
|  | 38. | 36 or 37 |
|  | 39. | ((scoping or systematic or critical or narrative or realist) adj3 review).tw,kf. |
|  | 40. | 35 and 38 and 39 |
| CINAHL |  | ((game or games) N3 (arcade? or console or consoles or computer* or digital* or internet* or mobile or offline or online or pc or video* or web or web-based or WWW or iphon* or "i-phon*" or smartphon* or "smart phon*" or android* or tablet* or Game boy or Game boys or Gameboy* or Gamecube or "N Gage" or "NES Classic" or Nintendo* or playstation* or play station* or "Super NES" or Wii or Xbox*)) OR ("call of duty*" or candy crush* or candycrush* or diablo* or "far cry*" or "first person shooter*" or "grand theft auto*" or "massively multi-player online*" or "massively multiplayer online*" or minecraft* or "mortal combat*" or "mortal kombat*" or "multi-player online battle arena*" or "multiplayer online battle arena*" or "pac man" or "pac manTM" or pokemon* or pong or sonic mania* or (sonic N2 hedgehog*) or starcraft* or "super mario*" or tetris* or titanfall* or tomb raider* or wolfenstein* or "world of warcraft*" or zelda*) OR ("online shopping" or "e-commerce" or "e-tail" or "e-shopping" or "delivery") OR (porn* or "sexually explicit material*" or SEM or erotic* or non-paraphilic or cyberporn* or cybersex or "sexual material" or "sexual content") OR ("social media" or "social network" or Facebook or LinkedIn or Twitter or Instagram or Pinterest or WeChat or Weibo or TikTok) AND ((addict* or compulsive* or detriment* or disorder* or excess* or habitual* or hazardous* or obsess* or problem* or pathologic* or risky) N5 behav*) OR ((addict* or compulsive* or depend* or detriment* or disorder* or excess* or habitual* or hazardous* or impair* or obsess* or overus* or (over N1 ("use" or used or uses or using)) or problem* or pathologic* or persist* or recur*) N5 (game or games or gaming or videogam*)) OR (habit or habits or habit-forming) AND (prevalen* or inciden* or epidemiolog* or burden) AND ((scoping or systematic or critical or narrative or realist) N3 review) |
| Cochrane | #1 | IGD |
|  | #2 | gam* or internet* or disorder* |
|  | #3 | #1 and #2 |

|  |  |
| --- | --- |
| #4 | [mh Video Games] |
| #5 | (game or games) NEAR/3 (arcade or console or consoles or computer* or digital* or internet* or mobile or offline or online or pc or video* or web or web-based or WWW or iphon* or i-phon* or smartphon* or (smart NEXT phon*) or android* or tablet* or Game boy or Game boys or Gameboy* or Gamecube or "N Gage" or "NES Classic" or Nintendo* or playstation* or (play NEXT station*) or "Super NES" or Wii or Xbox* or app or apps or Google Play or GooglePlay or iOS) |
| #6 | ((("call of" NEXT duty*) or (candy NEXT crush*) or candycrush* or diablo* or (far NEXT cry*) or ("first person" NEXT shooter*) or ("grand theft" NEXT auto*) or ("massively multi-player" NEXT online*) or ("massively multiplayer" NEXT online*) or minecraft* or (mortal NEXT combat*) or (mortal NEXT kombat*) or ("multi-player online battle" NEXT arena*) or ("multiplayer online battle" NEXT arena*) or "pac man" or "pac manTM" or pokemon* or pong or (sonic NEXT mania*) or (sonic NEAR/2 hedgehog*) or starcraft* or (super NEXT Mario*) or tetris* or titanfall* or (tomb NEXT raider*) or wolfenstein* or ("world of" NEXT warcraft*) or zelda*)) |
| #7 | ("online shopping" or "e-commerce" or "e-tail" or "e-shopping" or "delivery") |
| #8 | porn* or ("sexually explicit" NEXT material*) or SEM or erotic* or non-paraphilic or cyberporn* or cybersex or "sexual material" or "sexual content" |
| #9 | [mh "Sexual Behavior"] or [mh "Compulsive Sexual Behavior Disorder"] or [mh Erotica] |
| #10 | "social media" or "social network" or Facebook or LinkedIn or Twitter or Instagram or Pinterest or WeChat or Weibo or TikTok |
| #11 | [mh "Social Media"] |
| #12 | {OR #4-#11} |
| #13 | [mh "Behavior, Addictive"] or [mh Habits] or [mh "Health Behavior"] or [mh "Health Risk Behaviors"] or [mh "Social Behavior Disorders"] or [mh "Internet Addiction"] or [mh "Internet Addiction Disorder"] or [mh "Internet Gaming Disorder"] or [mh "Internet Usage"] or [mh "Technology Addiction"] or [mh "Video Game Addiction"] or [mh "Mobile Phone Addiction"] |
| #14 | (addict* NEAR/3 (computer* or digital* or internet or i-phon* or iphon* or online or smart phon* or smartphon* or video* or web or WWW)) |
| #15 | (addict* or compulsive* or detriment* or disorder* or excess* or habitual* or hazardous* or obsess* or problem* or pathologic* or risky) NEAR/5 behav* |
| #16 | (addict* or compulsive* or depend* or detriment* or disorder* or excess* or habitual* or hazardous* or impair* or obsess* or overus* or (over NEXT ("use" or used or uses or using)) or problem* or pathologic* or persist* or recur*) NEAR/5 (game or games or gaming or videogam*) |
| #17 | habit or habits or habit-forming |
| #18 | {OR #13-#17} |
| #19 | prevalen* or inciden* or epidemiolog* or burden |
| #20 | [mh Epidemiology] |
| #21 | {OR #19-#20} |
| #22 | (scoping or systematic or critical or narrative or realist) NEAR/3 review |
| #23 | ((#3 OR (#12 AND #18)) AND #21 AND #22) |

### 2.4. Data extraction and synthesis

We extracted author list, study title, digital object identifier, type of review, year of publication, number of studies included in the review, covered countries and regions, study population, time period of the review, type of problematic internet use, diagnostic criteria, prevalence rate, reported risk factors, and reported health outcomes from the included reviews, summarized in Table 2. One reviewer (SW) independently extracted data from the eligible reviews. A second reviewer (RVK) reviewed and validated the extraction. Data from individual reviews are showcased in tabulated format and synthesised narratively. AMSTAR quality assessment was performed by three independent reviewers (TST, GGB, ARU). Average scores on critical items were used to categorise reviews into “High,” “Moderate,” “Low,” and “Critically Low” confidence categories as defined by the creators of AMSTAR-2^21^. Inter-rater reliability between assessors was calculated using Krippendorff’s Alpha ^22^.

**Table 2:** Overview of the characteristics and key findings of the included literature reviews.

**Table 3:** Table of studies included in meta-analysis by region. EMR = Eastern Mediterranean Region, RA = Region of the Americas, EUR = European Region, SEAR = Southeast Asia Region, WPR = Western Pacific Region.

|  | Problematic Gaming | Problematic Internet Use | Problematic Smartphone Use | Problematic Social Media Use | Total |
| --- | --- | --- | --- | --- | --- |
| AFR | 1 | 4 | 2 | 3 | 10 |
| EMR | 8 | 39 | 14 | 16 | 77 |
| EUR | 74 | 139 | 18 | 27 | 258 |
| Mixed | 10 | 5 | 1 | 4 | 20 |
| RA | 12 | 20 | 9 | 4 | 45 |
| SEAR | 9 | 33 | 7 | 18 | 67 |
| WPR | 38 | 183 | 43 | 12 | 276 |
| Total | 152 | 423 | 94 | 84 | 753 |

### 2.5. Meta-analysis

Meta-analyses were performed for the following problematic internet use subtypes: gaming, internet, social media, and smartphone use. Sexting as described in Madigan et al. ^23^ was excluded because Madigan et al. ^23^ was the sole systematic review and meta-analysis of sexting prevalence, and cybersex addiction as described by Meng et al. ^8^ encompasses behaviors such as online pornography addiction that are distinct from sexting. Ostinelli et al. ^24^ was excluded from meta-analyses, as Darvesh et al.^25^ is a scoping review and Ostinelli et al. ^24^ does not estimate prevalence of problematic gaming. Prevalence estimates were manually extracted from Alimoradi et al.^26^ due to a lack of prevalence statistics in the original systematic review/meta-analysis.

#### 2.5.1 Overlapping studies

We first assessed overlap in studies included in each systematic review using corrected coverage area (CCA) as described in the literature^27^. Overlapping studies were manually identified and duplicates were removed. If prevalence estimates between overviews disagreed, primary literature was manually examined to assess the source of disagreement. Two main sources of disagreement were identified: (1) case definition for problematic internet users, (2) study population definition (e.g., general population versus internet users), and (3) stratification or lack thereof for primary studies containing multiple regional estimates. For primary studies where systematic reviews/meta-analyses identified conflicting numbers of cases, the prevalence estimate utilising a more liberal definition of problematic internet use was used. For instance, if one meta-analysis counted only those “severely addicted” to the internet and another counted the sum of “severely” and “moderately addicted,” the estimate from the latter was extracted to capture a broader spectrum of PUI. For primary studies with differing study populations between reviews, we took the more general population as opposed to—for example—restricting the sample size to internet users in the sample. There were three instances in which both reviews estimated prevalence from the same primary article, but one review divided estimates by region while another review aggregated estimates into one aggregate prevalence estimate. In these cases, we opted for the prevalence estimates stratified by region, as reported by the latter review.

#### 2.5.2 Random effects meta-analysis

We re-analysed each meta-analysis using extracted individual study prevalence estimates. Due to high heterogeneity in study design, population, and assessment tools, we used a random effects model with the double arcsine transformation^28^. We calculated I^2^ as a measure of between-study heterogeneity. Subgroup analysis was performed for region but not sex due to inconsistent reporting on male versus female cases in primary studies.

## 3. Results

Our search strategy yielded 1545 results (Medline: 187, Embase: 340, PsycInfo: 0, CINAHL: 348, Cochrane: 670) from academic database searches, 1421 (91.97%) after deduplication. Ultimately, we included 11 (0.71%) literature reviews. We found a crude inter-rater agreement score of between the two screeners (RVK and ARU) of 98.9%. We also accounted for the possibility of reaching inter-rater agreement by chance by computing Krippendorff’s Alpha (0.82), which points to strong reliability between the two screeners. The screening process is shown in the PRISMA flowchart in Figure 1. The 11 included review studies covered a total of 1,109 original research articles, representing a total of 3,145,428 individuals.

**Figure 1:**
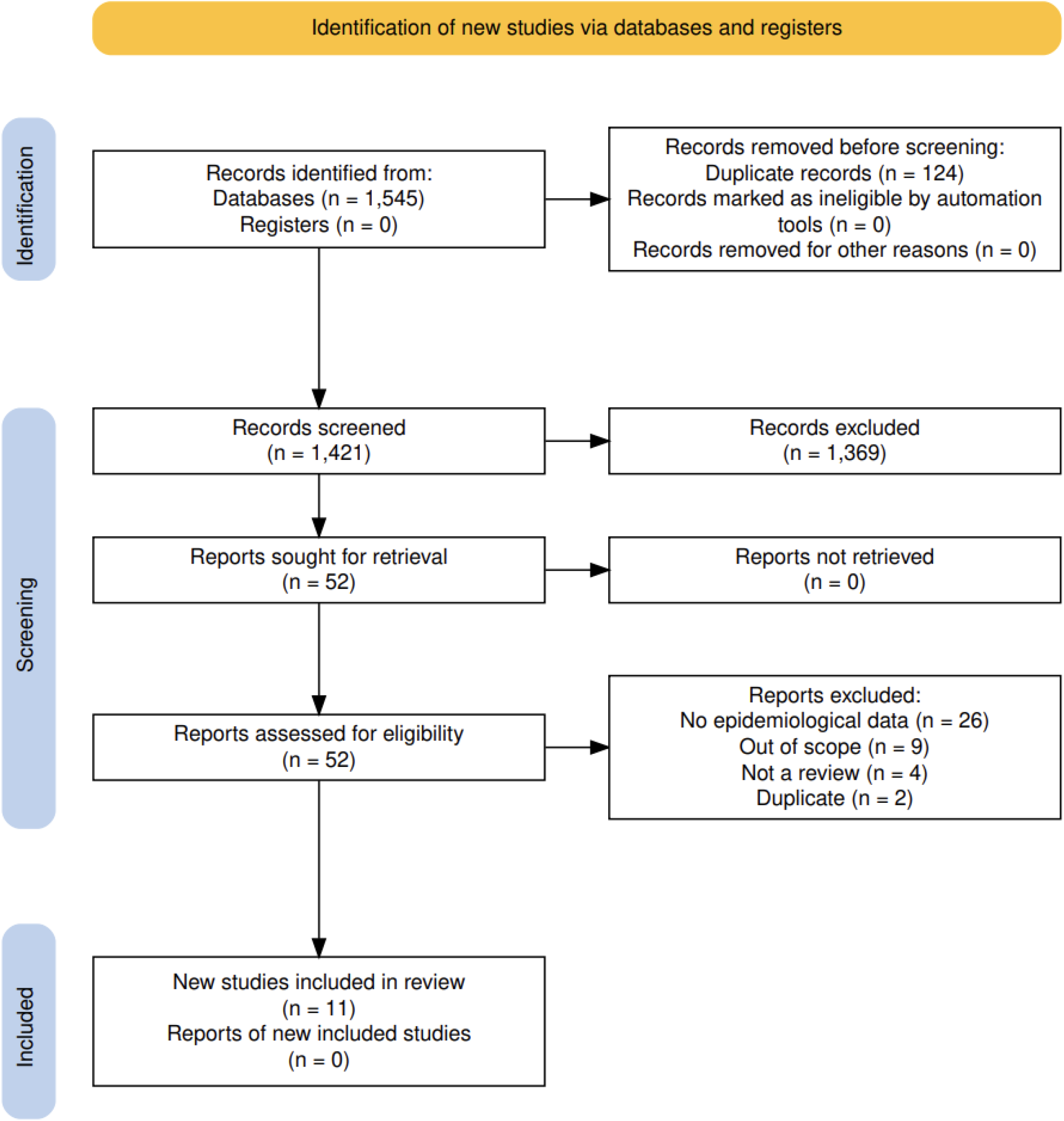
PRISMA flowchart of the screening process.

### 3.1 AMSTAR2 Quality Assessment

Critical appraisal suggests low confidence in PUI prevalence estimates. One systematic review fell into the “Moderate” category (Imperato et al.), five reviews fell into the “Low” category, and four reviews fell into the “Critically Low” category. The most common methodological flaws were in Criteria Item 7: including a list of excluded studies and justification for exclusion (missing in 70% of studies) and Criteria Item 2: a priori review method establishment and justification for significant deviations from the protocol (40% of studies). These results highlight a need for improved standards in transparency and reporting in systematic reviews and meta-analyses on PUI to improve confidence in prevalence estimates. We found a crude inter-rater agreement score of 81.8% between the three quality assessors (TST, GBB, and ARU). We also accounted for the possibility of reaching inter-rater agreement by chance by computing Krippendorff’s Alpha (0.77), which points to acceptable reliability between the three quality assessors.

### 3.2 Prevalence estimates from individual reviews

Details of review characteristics are summarised in Table 1. Ten of the included review studies were published in 2020 or later^8,21,24,26,28–33^. Ten of the included reviews were global in scope^8,21,23,24,26,28–31^, while one focused on PUI in Latin America and the Caribbean^28^. Study populations vary widely, ranging from specific cohorts (e.g., 110,380 individuals under 18 years old ^23^ or 35,520 university students^26^) to population-level cohorts spanning between 17 and 64 countries ^8,20,24–26,28–30^. One review did not report on their study population in scope ^31^. The timeframes captured by the included reviews were also variable and ranged from 1990 to 2022. In terms of types of PUI being covered by the included reviews, seven reviews focused on some variation of gaming disorder or addiction ^20,24,25,28–31^, three reviews on digital or internet addiction as an umbrella ^8,20,26^, one on social media addiction ^34^, and one on sexting^23^. Notably, only one of the included reviews restricted their methodology to any specific diagnostic criteria, namely the Lemmens Internet Gaming Disorder-9 Scale ^29^. Another review mapped the diagnostic criteria used in their included studies, which comprised the DSM-5 Internet Gaming Disorder scale, a self-reported instrument based on DSM-5 Internet Gaming Disorder criteria, the Gaming Addiction Scale, the Ten-item Gaming Disorder Test (IGDT-10), and the Brazilian version of the Internet Gaming Disorder Scale-Short-Form (IGDS9-SF)^33^.

The prevalence rates of PUI vary across different sub-types and regions. Internet addiction in general showed a prevalence of 10.6% in one review ^26^, 14.22% (95% CI, 12.90%–15.65%) in a second review ^8^, and 7.02% (95 % CI, 6.09 %–8.08 %) in a third ^20^. The prevalence of sending sexually explicit content was estimated at 14.8% (95% CI, 12.8%-16.8%) with consent and 12.0% (95% CI, 8.4%-15.6%) without consent, while receiving was estimated at 27.4% (95% CI, 23.1%-31.7%) with consent and 8.4% (95% CI, 4.7%-12.0%) without consent^23^. Social media addiction was reported at a prevalence rate of 18.4% globally (95% CI, 14.7%-22.6%), though regional variations were also highlighted with Asia reporting a prevalence rate of 22.8% (95% CI, 18.5%-27.6%), Europe of 12.4% (95% CI, 5.3%-26.6%), Africa at 9.6% (95% CI, 1.1%-50.5%), and America at 15.8% (95% CI, 4.8%-41.1%) ^34^. Another study estimated social media addiction at 15.1% ^26^, while a third reported a prevalence of 17.42% (95% CI, 12.42%– 23.89%) ^8^. Smartphone addiction was estimated in one review at a prevalence rate of 26.99% (95% CI, 22.73%–31.73%) ^8^, and at 30.7% in a second review ^26^.

Gaming addiction, being covered most extensively by the included studies, showed a smaller variance in its reported prevalence rates, generally ranging between 2.47% (95% CI, 1.46 %– 4.16 %) and 6.04% (95% CI, 4.80%–7.57%)^8,21,28,29,31^. However, two studies reported particularly wide prevalence ranges. Hernandez-Vasquez and colleagues reported a prevalence range for gaming addiction between 1.1% and 38.2%, though the authors also explain that the high variability of instruments to define gaming disorder used in the different studies demonstrates that the prevalence rates and their interpretation should be handled with caution^33,35^.

### 3.3 Risk factors

Risk factors for PUI varied by behaviour type, demographic, and regional context. For sexting, prevalence reportedly increased with age and is higher among males ^23^. Social media addiction shows a temporal trend, with prevalence rising over time, as well as a gender bias with males being at higher risk ^26^. Digital addictions were more common among males, older individuals, and people from low- and middle-income countries ^8,31^. Risk factors for gaming addiction were older age, male gender, emotional dependence, social detachment, frequent and prolonged gaming sessions, and being in a state of increased emotional and psychological distress ^28,29,31^. Finally, health outcomes were only listed for gaming addiction, including lower academic scores, mental health problems such as depression, anxiety, and other addictive behaviors, and decreased self-esteem, life satisfaction, and social support ^31,36^.

**[insert PUI_Manuscript_Table2.docx]**

### 3.4 Meta-analysis

#### 3.4.1 Study overlap

Overlap between reviews was low for problematic gaming (4.9%), social media use (4.7%), and smartphone use (0%). Overlap was moderate for problematic internet use (8.4%). Although overlap fell in the moderate range for problematic internet use, there was a sizeable overlap between Pan et al. and Meng et al. (52.6% of primary studies in Pan et al. are also in Meng et al.) and between Alimoradi et al. and Meng et al. (24.5% of primary studies in Alimoradi et al. are also in Meng et al.) [8, 21, 30].

#### 3.4.2 Pooled prevalence estimates

Forest plots for each PUI subtype are reported, along with funnel plots (Fig. 2-6). Across all regions, pooled prevalence for problematic gaming is estimated at 6% [CI: 5-7%], pooled prevalence for problematic internet use is estimated at 16% [CI: 15-17%], pooled prevalence for problematic smartphone use is estimated at 32% [CI: 28-35%], and pooled prevalence for problematic social media use is estimated at 23% [CI:19-28%]. Across regions and PUI subtypes, there is high study heterogeneity as indicated by I^2^>99% for every PUI subtype. There is evidence of funnel plot asymmetry across PUI subtypes, suggestive of small study bias.

**Figure 2:**
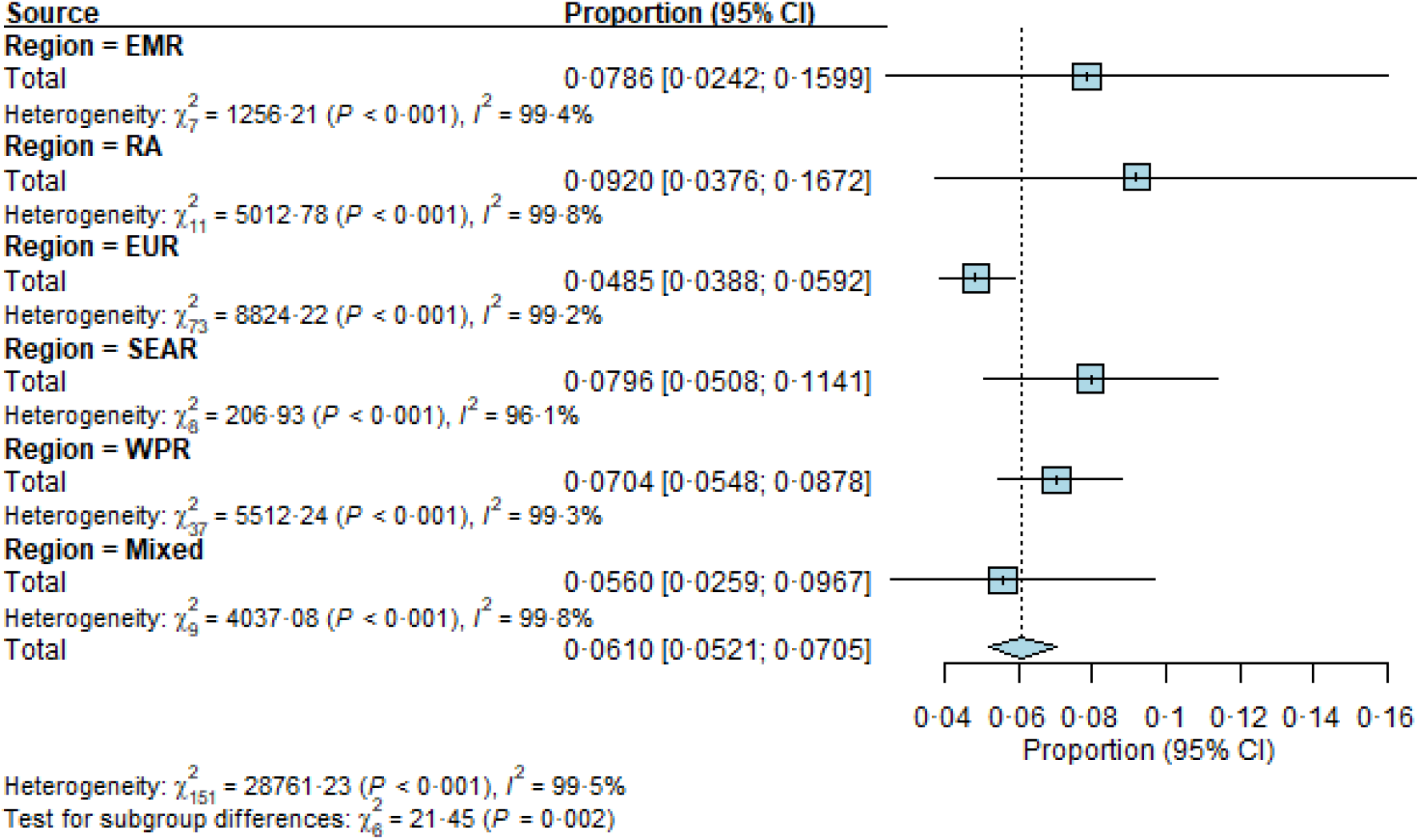
Problematic gaming by WHO region. EMR = Eastern Mediterranean Region, RA = Region of the Americas, EUR = European Region, SEAR = Southeast Asia Region, WPR = Western Pacific Region.

**Figure 3:**
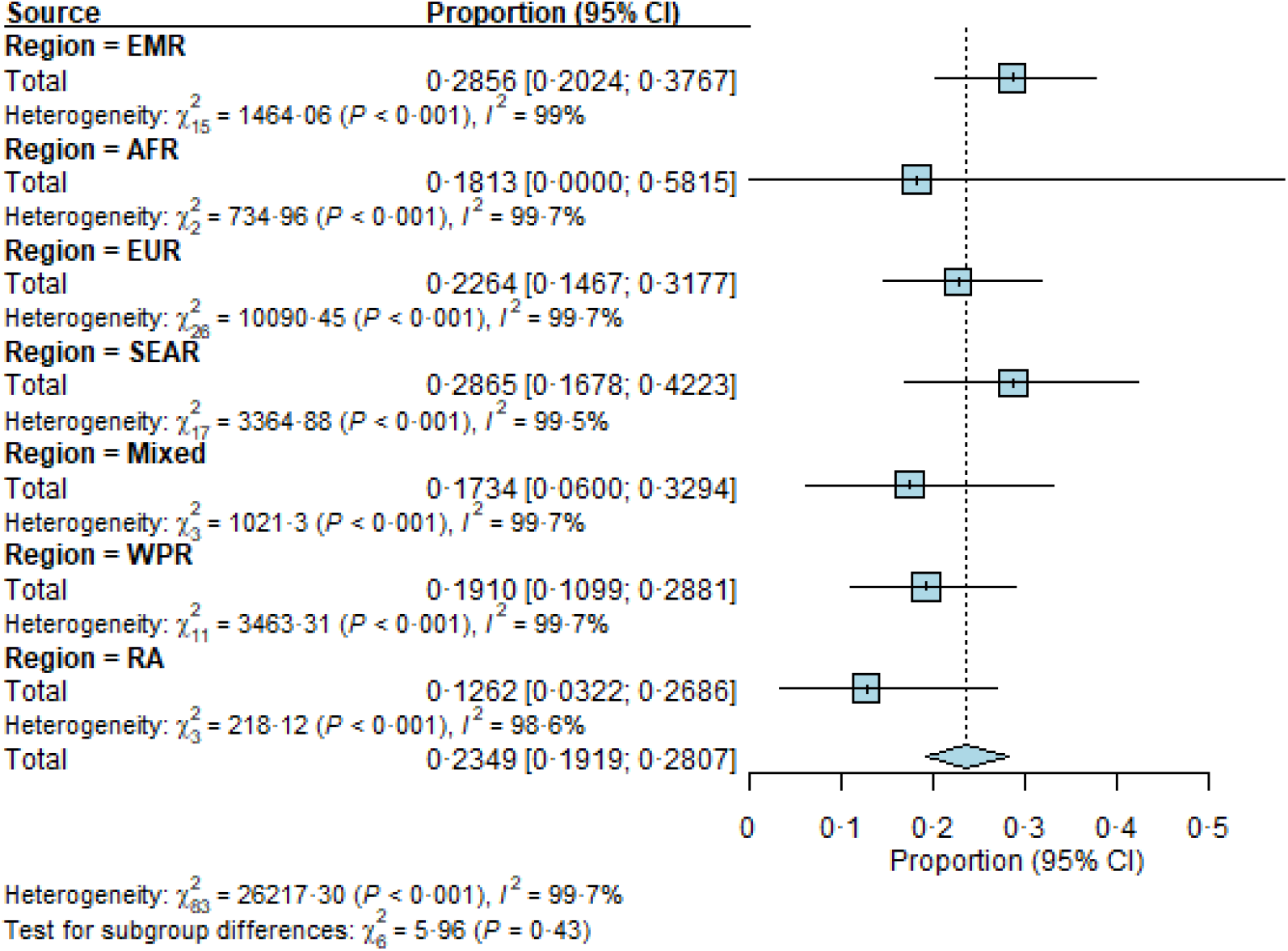
Problematic social media use by WHO region. EMR = Eastern Mediterranean Region, RA = Region of the Americas, EUR = European Region, SEAR = Southeast Asia Region, WPR = Western Pacific Region.

**Figure 4:**
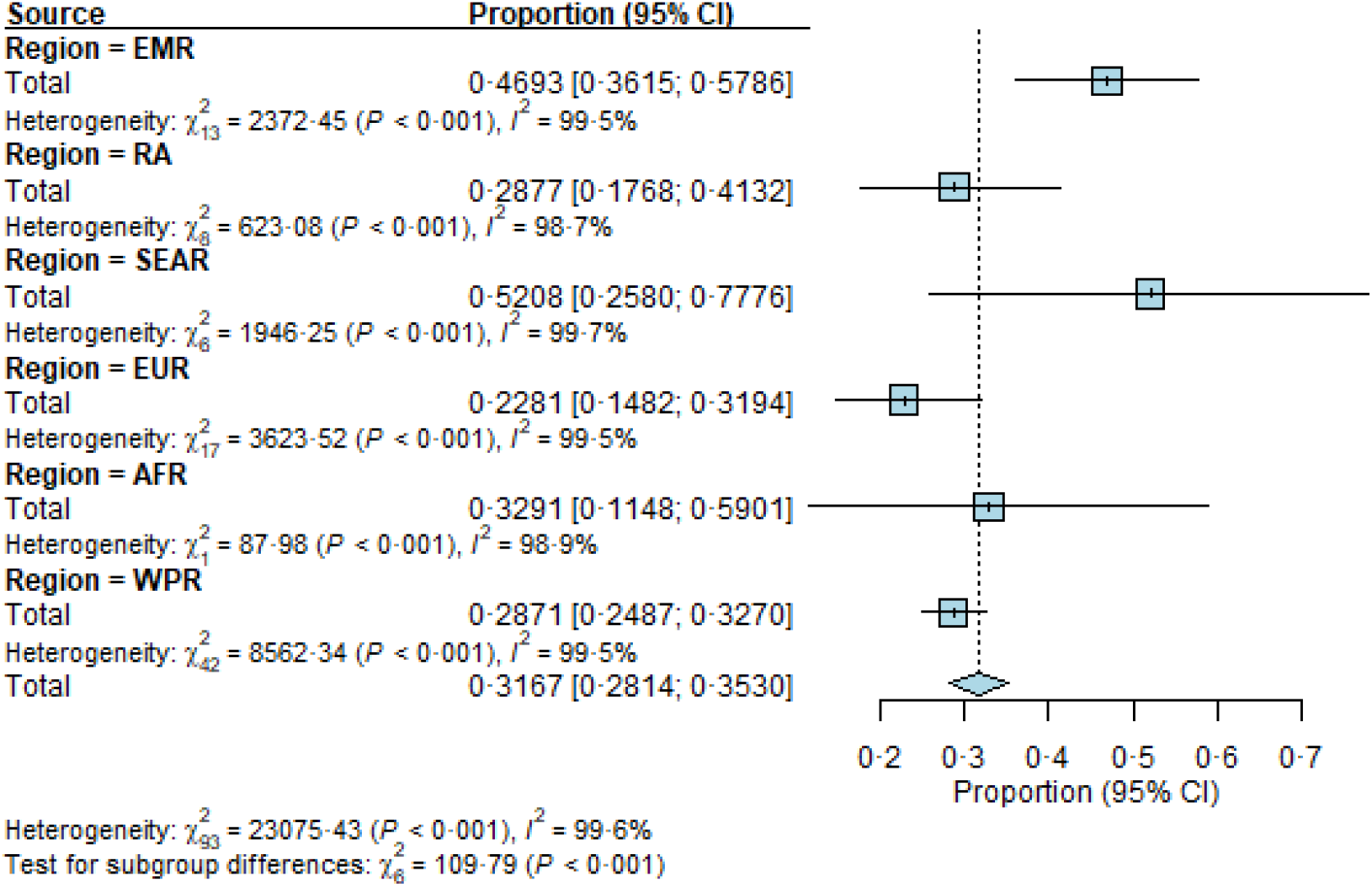
Problematic smartphone use by WHO region. EMR = Eastern Mediterranean Region, RA = Region of the Americas, EUR = European Region, SEAR = Southeast Asia Region, WPR = Western Pacific Region.

**Figure 5:**
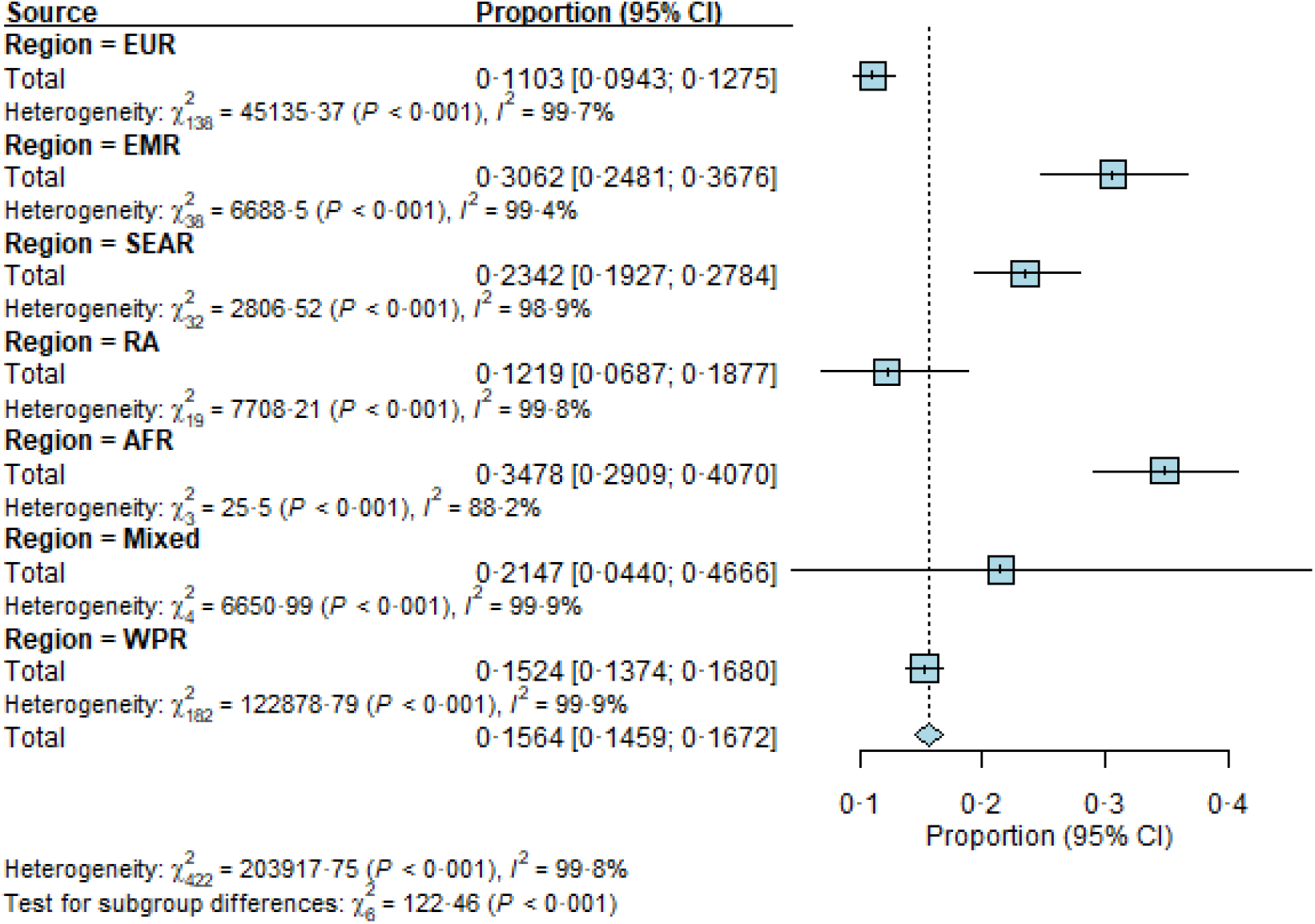
Problematic internet use by WHO region. EMR = Eastern Mediterranean Region, RA = Region of the Americas, EUR = European Region, SEAR = Southeast Asia Region, WPR = Western Pacific Region.

**Figure 6:**
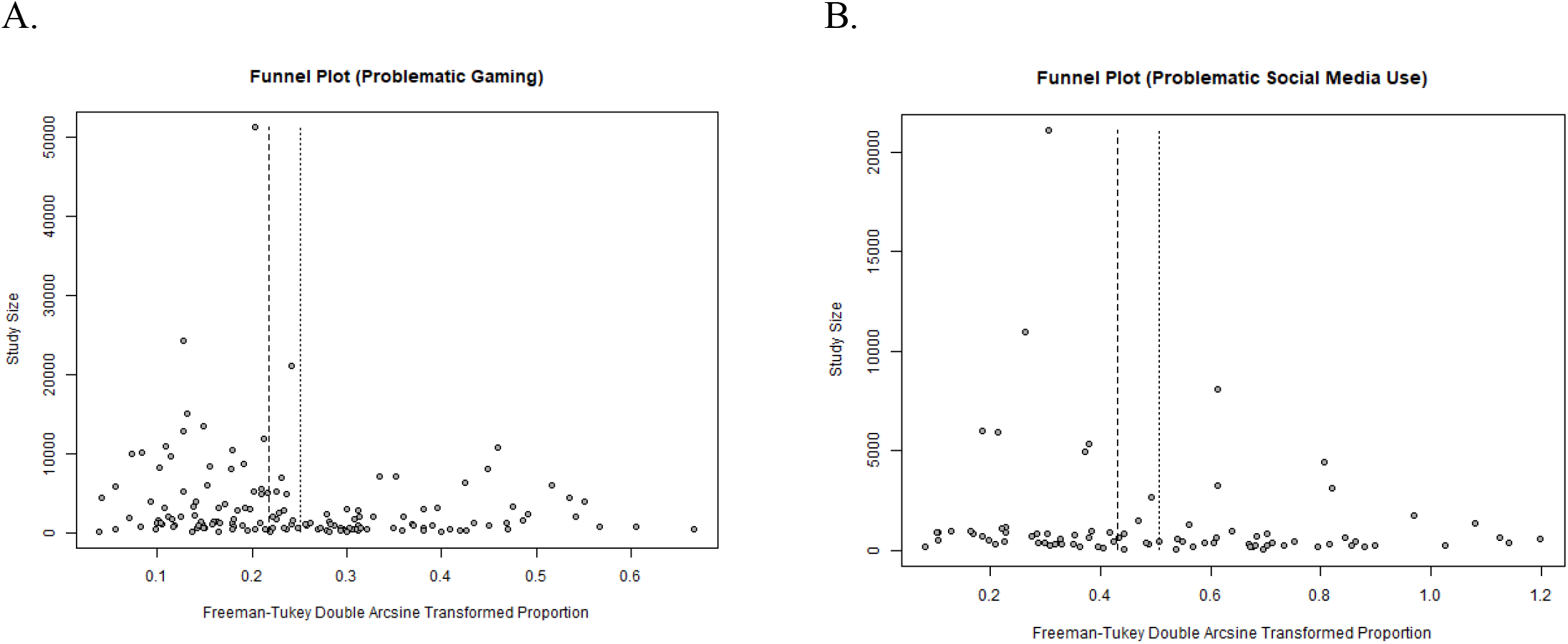
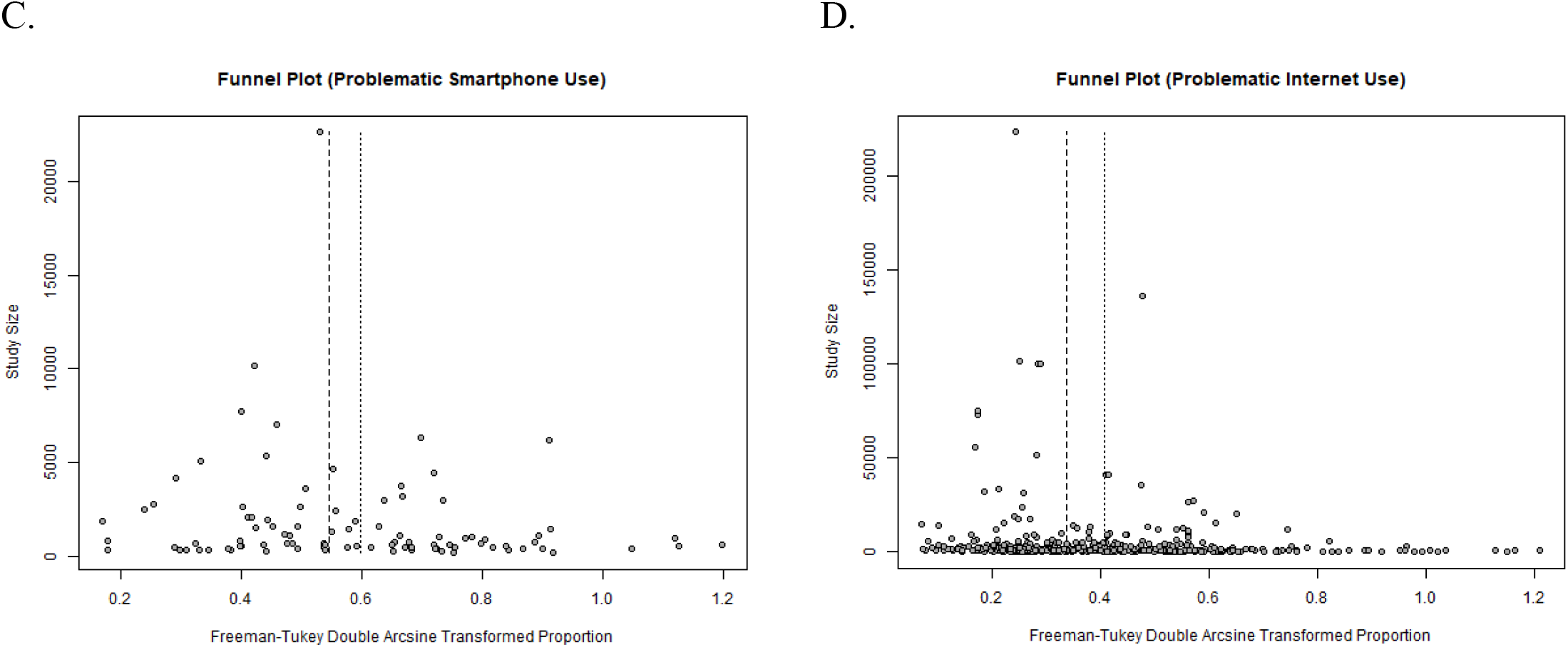
Funnel plots for each meta-analysis by subtype. Thick vertical dashed line represents common effects estimate, thin vertical dotted line represents random effects estimate.

#### 3.4.3 Regional variation

The number of studies per region included in the meta-analysis for each subtype are reported in Table 2. Most studies across subtypes were conducted in the European and Western Pacific regions, with the lowest number of studies from the African region. Problematic gaming prevalence is highest in the Region of the Americas (9%, [CI: 4-17%]). Problematic social media use prevalence is highest in the Eastern Mediterranean (29% [CI: 20-38%]) and Southeast Asian regions (29% [CI: 17-42%]). Problematic smartphone use is most prevalent in the Southeast Asian region (52% [CI: 26-78%]). Estimates for general problematic internet use were highest in the African region (35% [CI: 29-41%]), however this estimate must be interpreted with caution given only four studies reported from the region on internet use specifically and 10 studies encompassing different PUI behaviours.

## 4. Discussion

This article was, to the best of our knowledge, the first comprehensive umbrella review of evidence synthesis works related to the epidemiology of PUI. Our review encompasses evidence syntheses covering five broad categories of PUI in a global context: sexting, general problematic internet use, problematic gaming, problematic social media use, and problematic smartphone use. Altogether, studies suggest a considerable proportion of the global population is affected by some form of PUI, ranging from 6% [CI: 5-7%] affected by problematic gaming to 32% [CI: 28-35%] affected by problematic smartphone use. Common risk factors for digital addictions across subtypes include older age, male gender, and psychological distress. Within the scope of our literature review search strategies, we only found evidence synthesis on the outcomes of problematic gaming in particular; this analysis reveals several consequences of problematic gaming, specifically lowered academic performance and increased risk of other mental health disorders and addiction behaviours. That said, there are important methodological issues across evidence syntheses that limit confidence in these findings. Most reviews included fall in the “Low” or “Critically Low” confidence categories as dictated by AMSTAR2 critical appraisal.

This umbrella review reveals several important sources of uncertainty around current epidemiological findings on PUI, namely disparities in research conducted on different PUI subcategories, substantial heterogeneity in study design and populations, and fragmented operationalisations of PUI. Our findings highlight that – despite a comprehensive search string covering multiple aspects of PUI, research on the prevalence of PUI is heavily concentrated in the field of gaming addiction and behaviours with some focus on social media and general digital addictive behaviours. This is alarming, since our results equally indicate that different tropes of PUI are associated with vastly different prevalence estimates. It is worth noting that, while gaming addiction and its various conceptualizations are comparatively heavily researched, their prevalence rates generally range between the 3% and 5% of the population, indicating a lower burden of disease compared to the prevalence rates of other tropes of PUI that range anywhere between the 10% and 30%. In contrast, research on the epidemiology of problematic use of social media, smartphones, or pornography remains scarce, even though extensive research has been performed in these topic areas in terms of clarifying risk groups, risk factors, and associations with a range of health determinants ^33,35,37,38^. Finally, at the regional level, despite existing research suggesting that those in lower- and lower-middle income countries are disproportionately affected by PUI, there is scarcity of research in the Americas, Southeast Asia, and Africa surrounding PUI^8,23^.

In addition to regional variation, there is substantial variation in study design. The most notable areas of ambiguity lie in the specific study populations and the tools and cut-offs used to assess internet use. Regarding study population, all the systematic reviews/meta-analyses incorporated primary studies of specific subpopulations that could have different risks of PUI compared to the general population, such as clinical populations, gamers, students, and Internet users. Furthermore, numerous studies were conducted during the COVID-19 pandemic, with many primary articles published around 2020 conducting their research online. Most reviews did not explicitly report or restrict their syntheses to specific forms of sampling. While most reviews did not restrict age or sex characteristics in the interest of investigating PUI in the general population, they often included primary studies using snowball or convenience sampling strategies that compromise the generalisability of their findings to the general population. We were not able to generate aggregate estimates for age- or sex-based subgroups because (1) evidence syntheses frequently only provided summary-level information on age and sex composition of primary studies, and (2) primary studies frequently did not report on prevalence stratified on these characteristics. The heterogeneity in study population underscores a need for PUI assessment in more robust, representative samples to obtain clearer prevalence estimates.

As for tools, there is not only a wide range of tools used to assess each PUI subtype, but a wide range of cutoffs and definitions used to categorize internet users. Over 50 different tools were used across primary studies included in the systematic reviews/meta-analyses, highlighting a need to validate and streamline tools of investigation. Further work is also necessary to establish clear classifications of PUI. For instance, in the case of general problematic internet use, some primary articles use a binary classification as addicted or not addicted to the internet, while others operationalise problematic internet use as “healthy,” “problematic,” and “pathological”^8,20^. Disagreements in prevalence estimates extracted from such primary articles often depend on whether the authors decided to sum several categories of problematic use or only count the most severe category (“pathological” or “severely addicted”). These differences in prevalence extraction highlight a need to disambiguate the terms “internet addiction,” “problematic use of the internet,” and “pathological use of the internet” to define the burden of PUI and its various manifestations more clearly.

## 5. Conclusion

Our umbrella review suggests that PUI behaviours likely affect a sizable proportion of the global population, but existing primary literature and, consequently, evidence syntheses suffer from numerous non-negligible methodological limitations. In light of rapid digital transformation, it is crucial that policy developments are informed by a foundation of high-quality epidemiological evidence grounded in clear PUI definitions^6^. Given the gaps and methodological issues we have identified in the literature, we propose several priorities for establishing a more robust evidence base. Across the board, work is needed to clarify the conceptual framework underlying PUI and which tools are optimal for measuring PUI. Regarding specific behaviours, we suggest prioritisation of internet use behaviours outside of problematic gaming and general problematic internet use, which were the most extensively researched in the literature we surveyed. In terms of region, research on PUI behaviours in lower- and lower-middle income countries is urgently needed. Finally, since many existing evidence syntheses—ours included—aggregate prevalence estimates from a mixture of school-based, clinical, and general population samples, more high-quality primary studies are needed to characterise disparities in PUI between specific subgroups of the population such that public health initiatives can prioritise those at highest risk of developing PUI and its concomitant health consequences. Significant refinement of epidemiological research on PUI with standardised measurement and nomenclature is urgently needed to appropriately prioritise and develop interventions for this increasing pressing digital public health issue.

## Data Availability

All data is in the public domain

## Acknowledgements

This work was funded by the European Union (BootStRaP project grant number: 101080238). Views and opinions expressed are however those of the author(s) only and do not necessarily reflect those of the European Union or the European Health and Digital Executive Agency (HaDEA). Neither the European Union nor the granting authority can be held responsible for them.

This work was supported by UK Research and Innovation program (the University of Cambridge project number: 10077671, the Euro Youth Mental Health project number: 10082693, the University of Hertfordshire project number: 10075008, the University of Southampton project number: 10075760). UK Research and Innovation accepts no liability, financial or otherwise, for expenditure or liability arising from the research funded by the Grant except as set out in these Terms and Conditions, or otherwise agreed in writing.

This work has received funding from the Swiss State Secretariat for Education, Research and Innovation (SERI).

---

**Figure 1:**
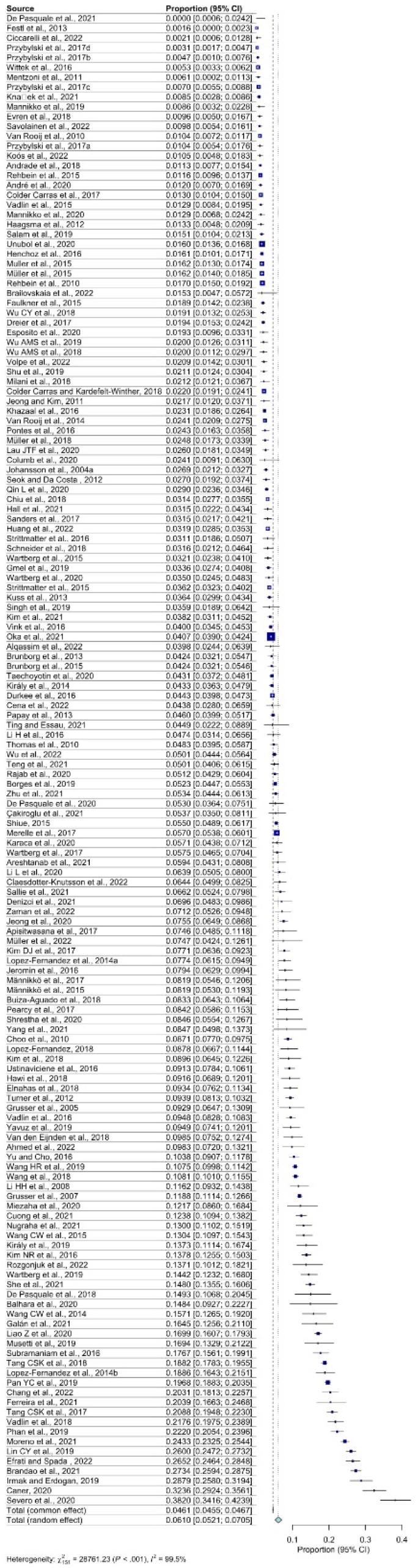
Problematic gaming - author forest plot.

**Figure 2:**
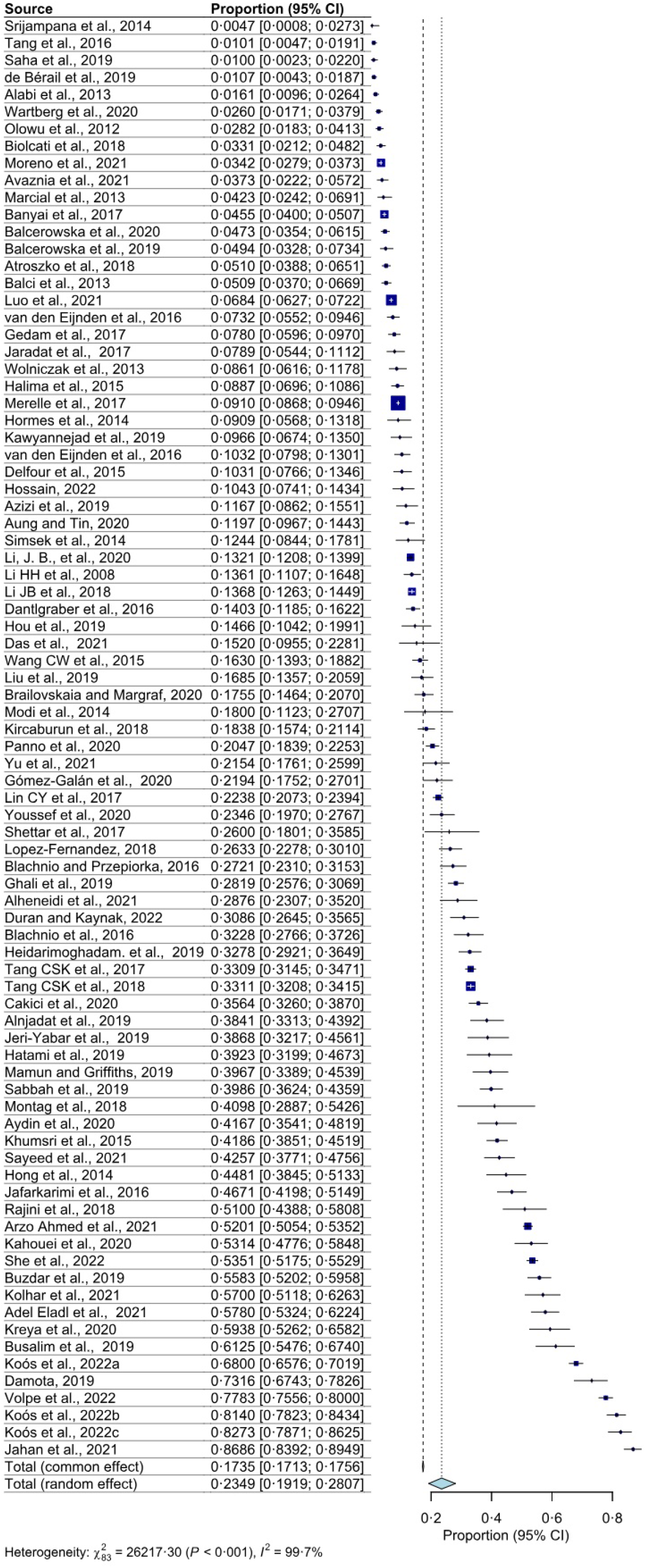
Problematic social media use - author forest plot.

**Figure 3:**
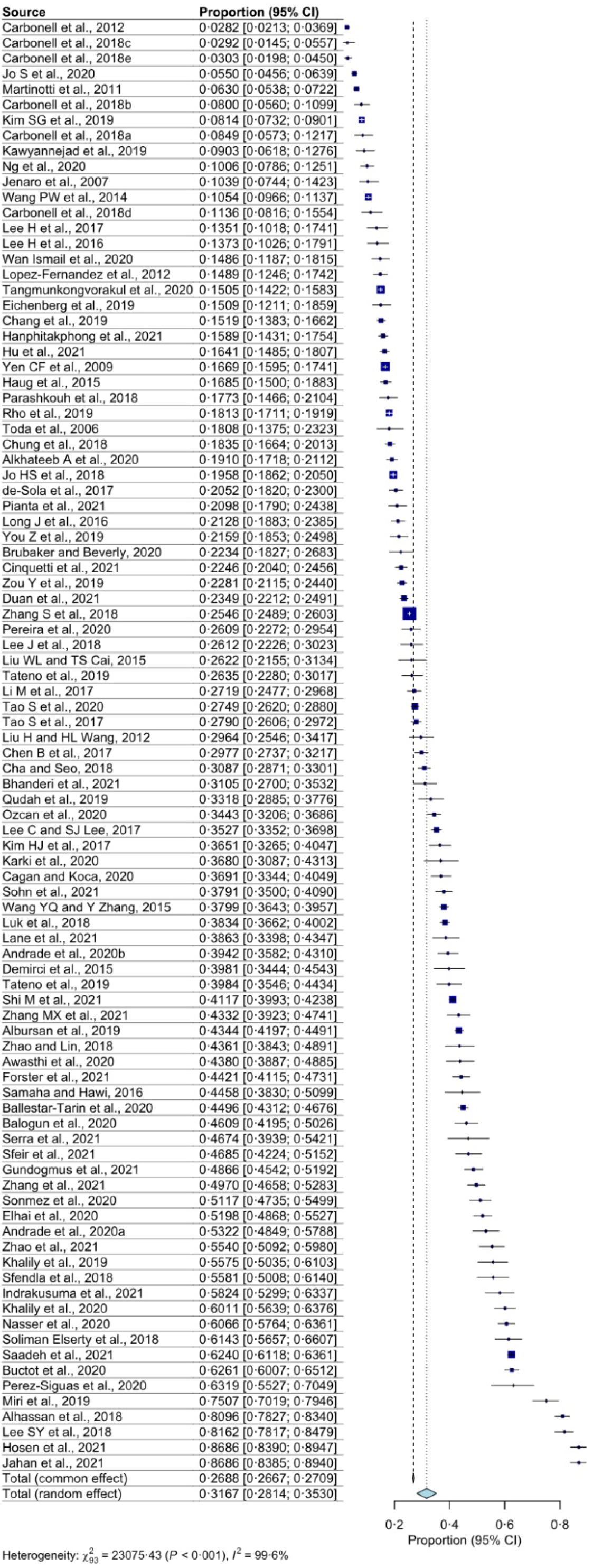
Problematic smartphone use - author forest plot.

**Figure 4:**
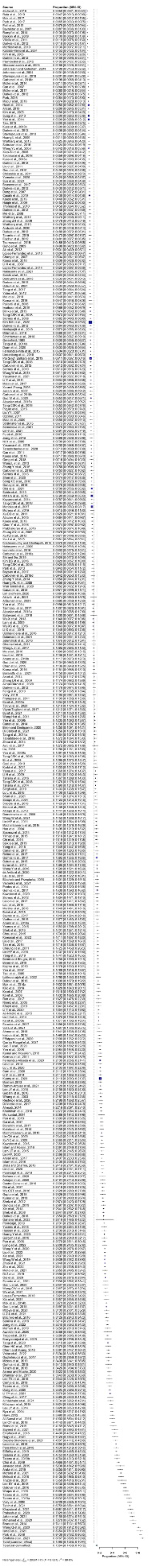
Problematic internet use - author forest plot.

**Figure 5:**
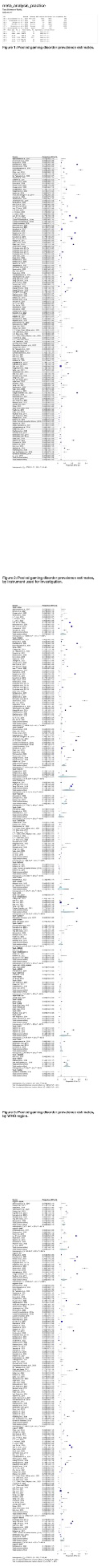
Meta-analysis figures.

**Figure 6:**
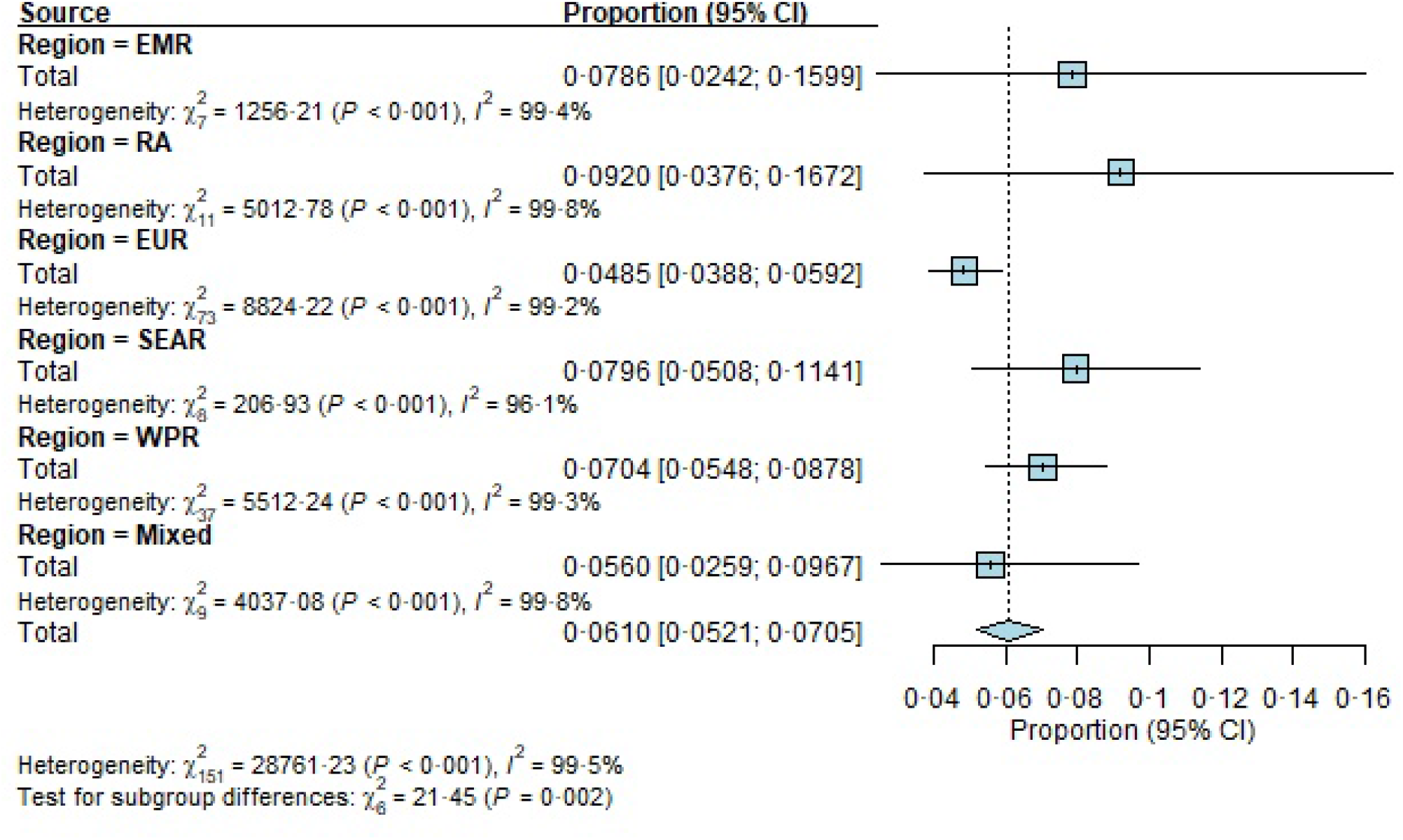
Gaming - regional subgroup forest plot.

**Figure 7:**
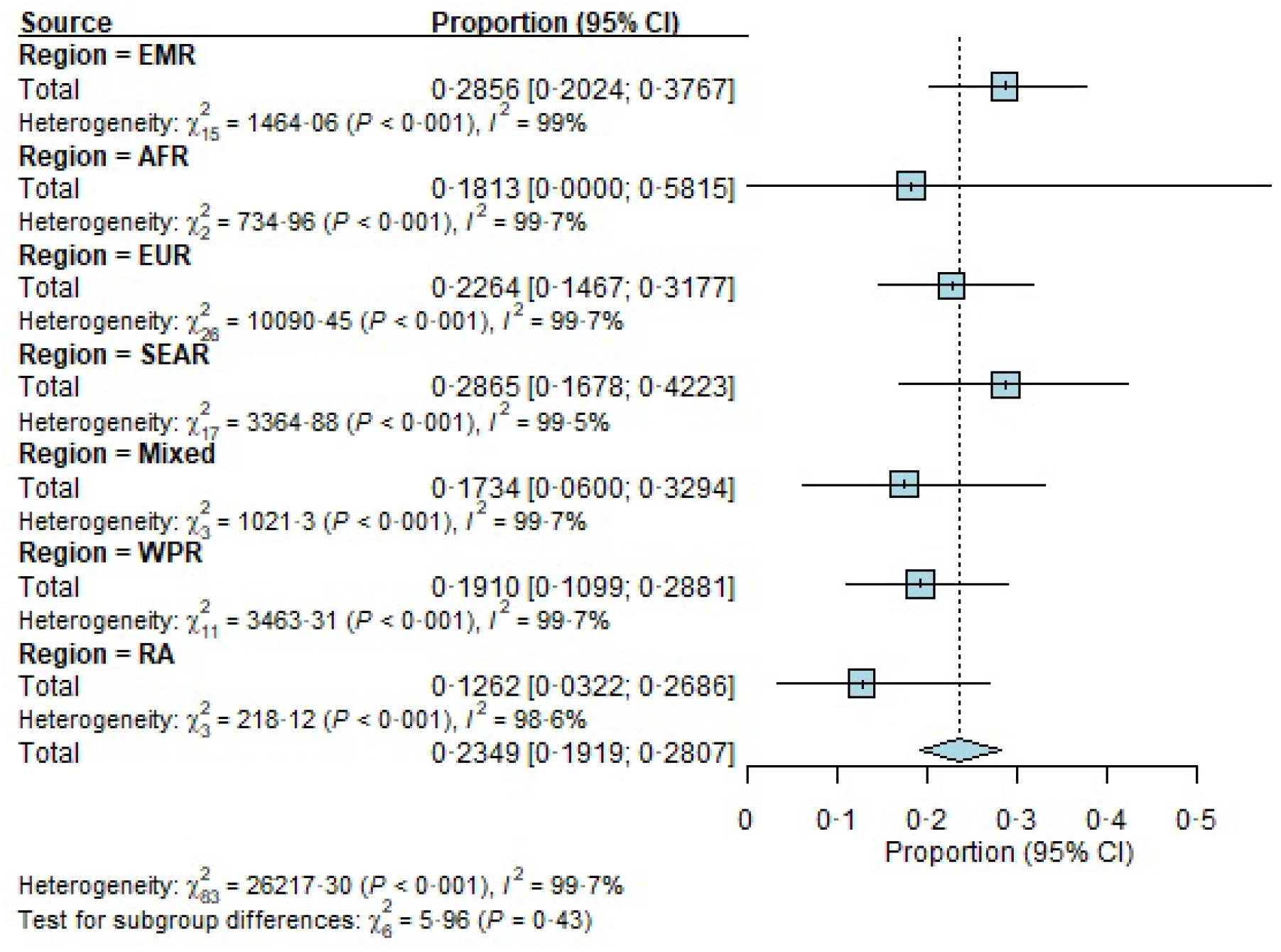
Social media - regional subgroup forest plot.

**Figure 8:**
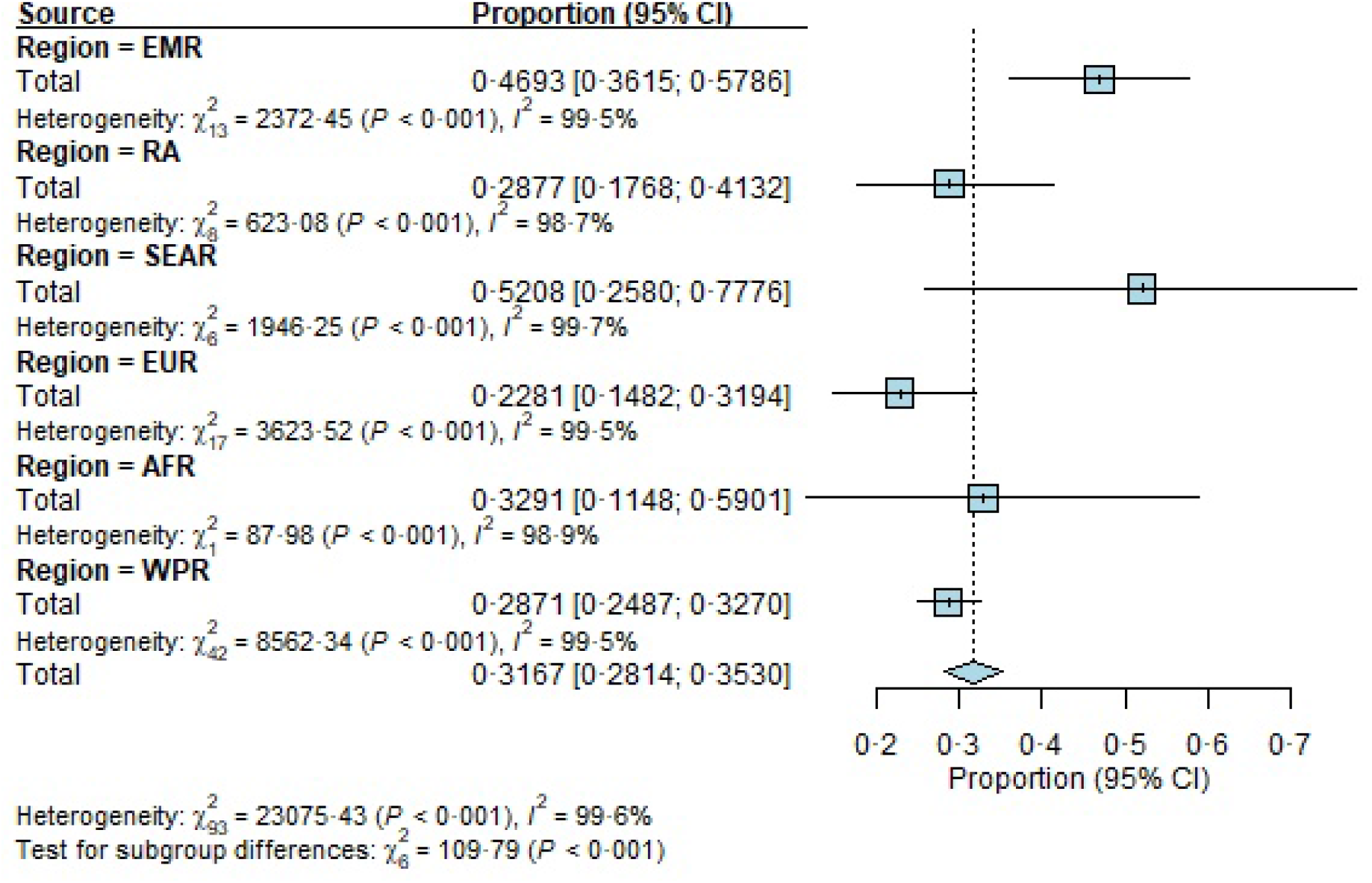
Smartphone - regional subgroup forest plot.

**Figure 9:**
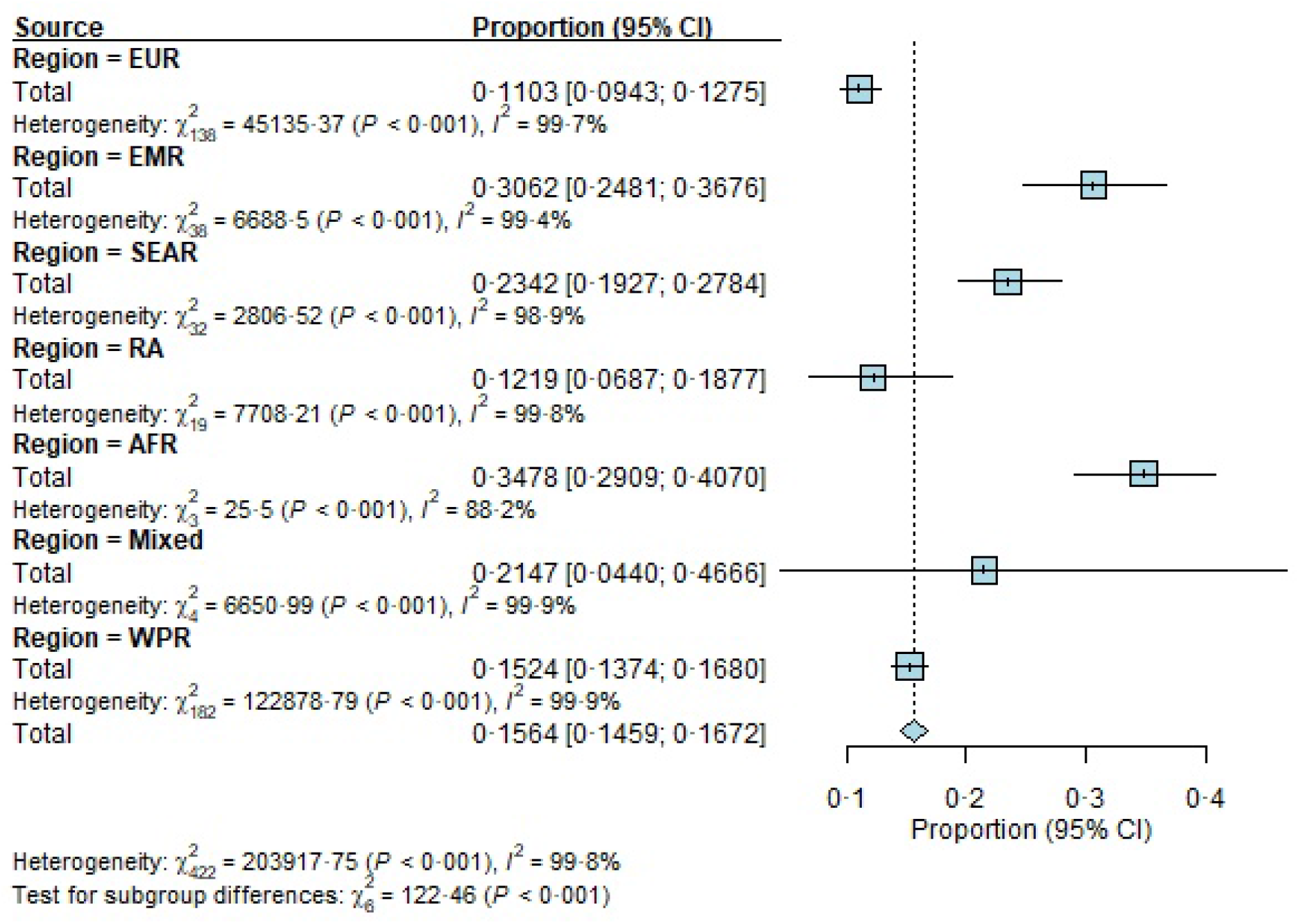
Internet - regional subgroup forest plot.

**Figure 10:**
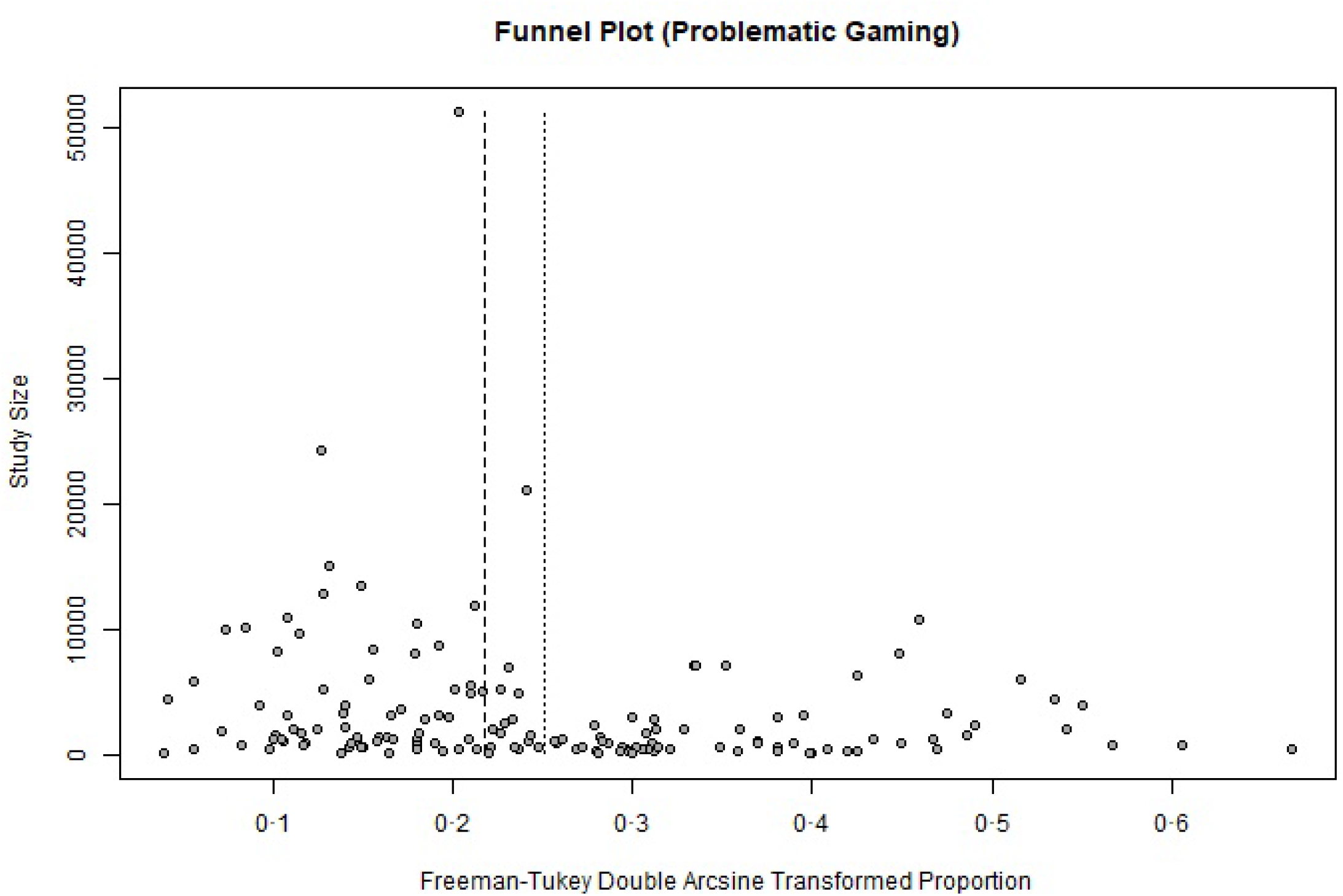
Gaming - funnel plot.

**Figure 11:**
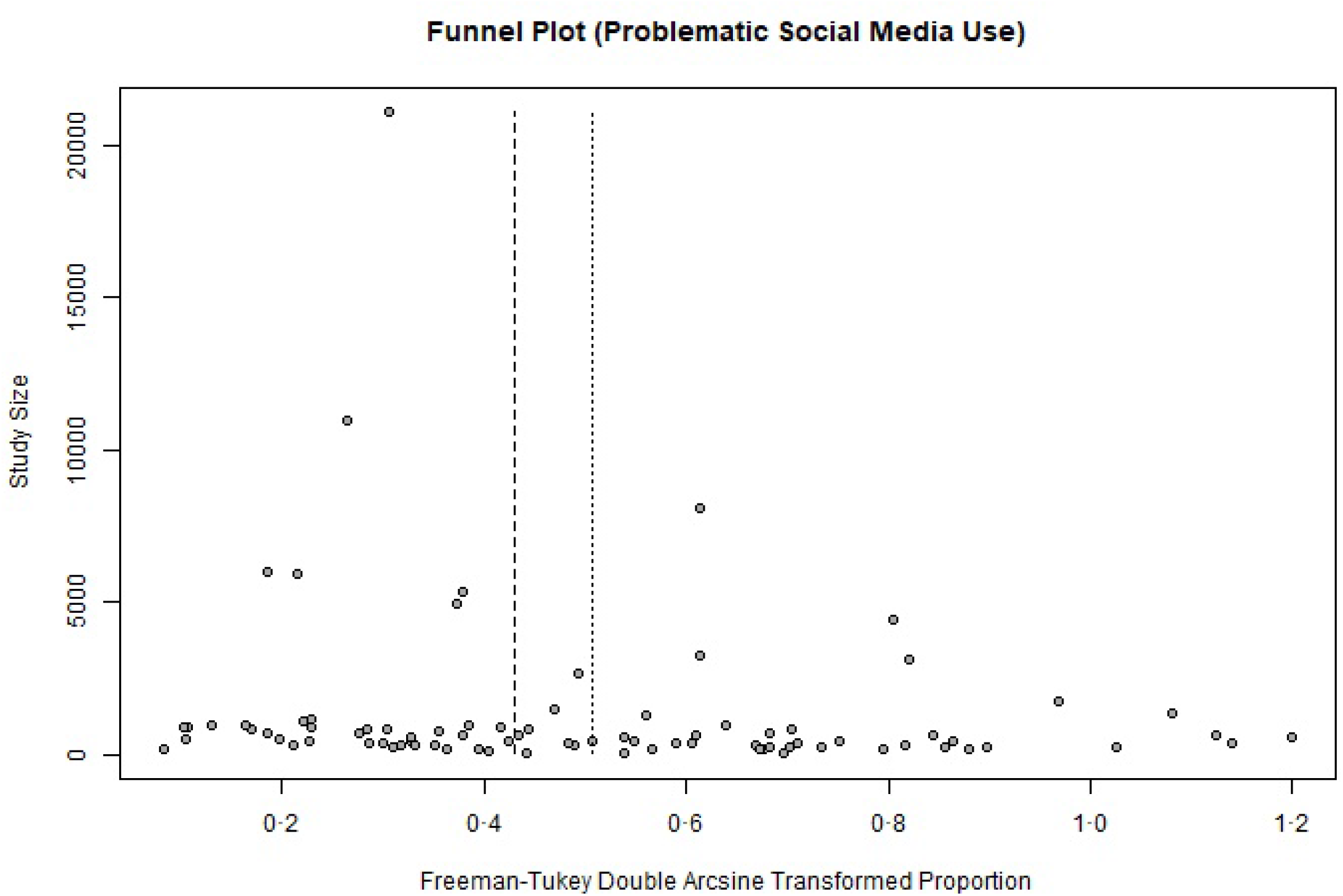
Social media - funnel plot.

**Figure 12:**
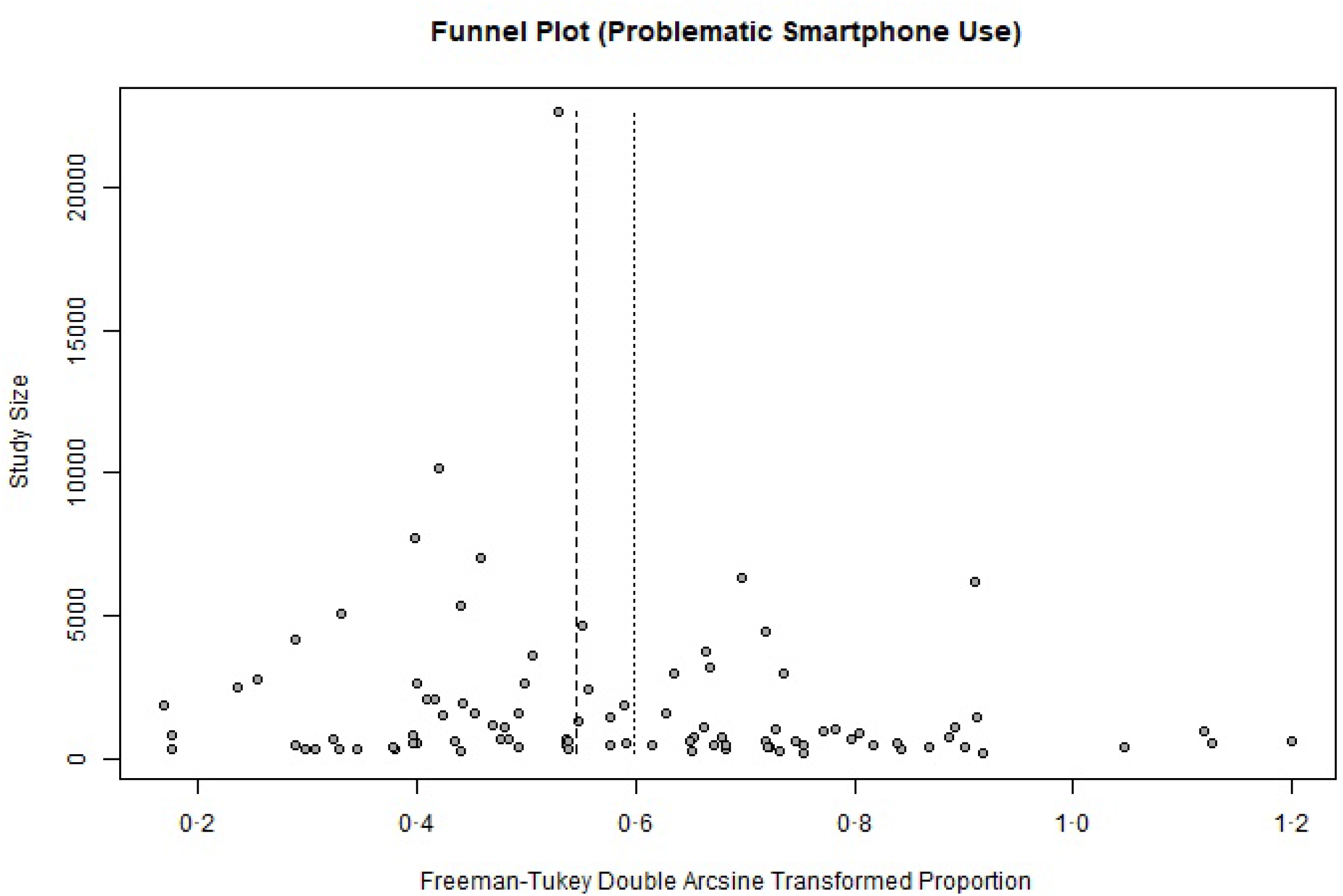
Smartphone - funnel plot.

**Figure 13:**
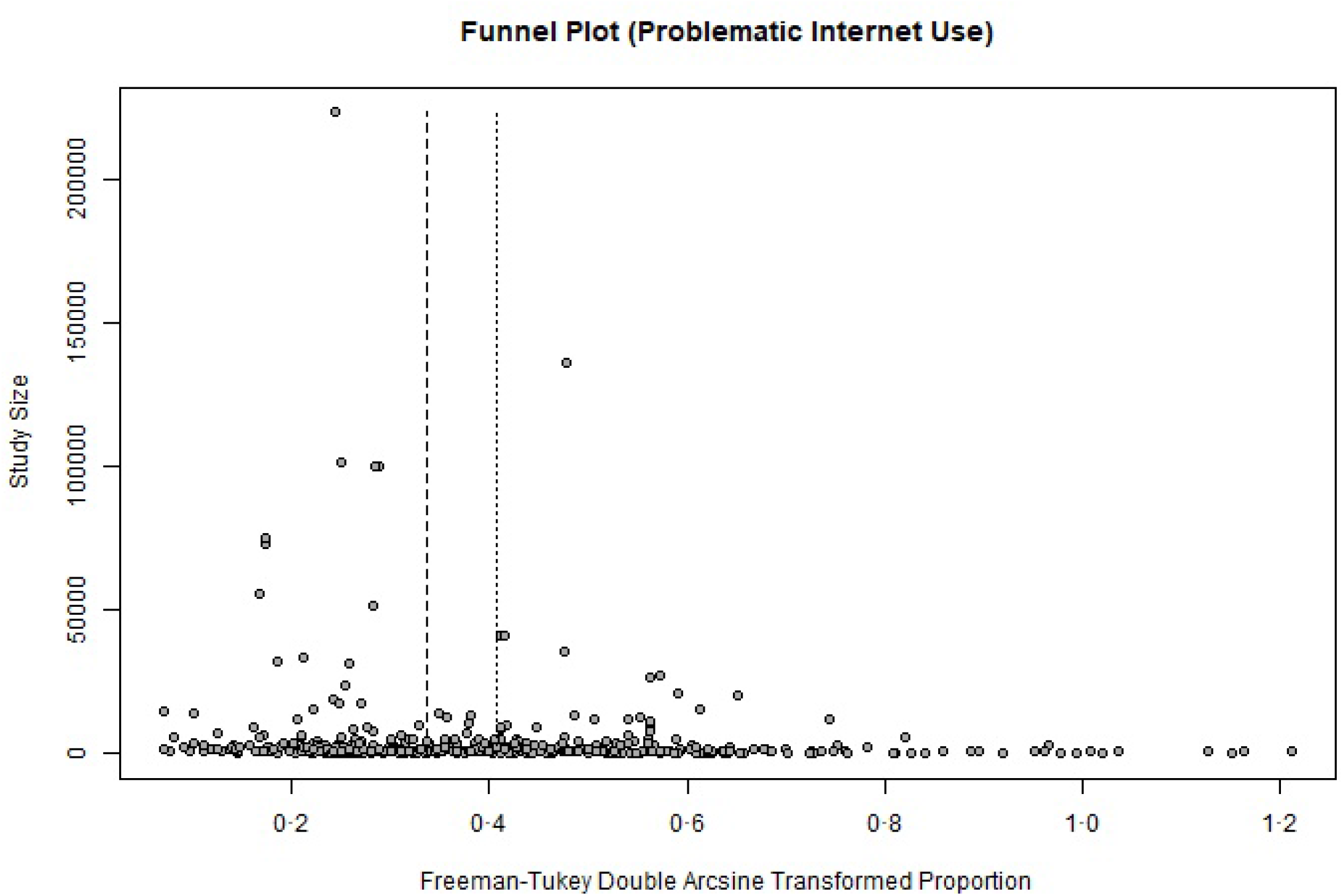
Internet - funnel plot.

